# Causal and Associational Language in Observational Health Research: A systematic evaluation

**DOI:** 10.1101/2021.08.25.21262631

**Authors:** Noah A. Haber, Sarah E. Wieten, Julia M. Rohrer, Onyebuchi A. Arah, Peter W.G. Tennant, Elizabeth A. Stuart, Eleanor J. Murray, Sophie Pilleron, Sze Tung Lam, Emily Riederer, Sarah Jane Howcutt, Alison E. Simmons, Clémence Leyrat, Philipp Schoenegger, Anna Booman, Mi-Suk Kang Dufour, Ashley L. O’Donoghue, Rebekah Baglini, Stefanie Do, Mari De La Rosa Takashima, Thomas Rhys Evans, Daloha Rodriguez-Molina, Taym M. Alsalti, Daniel J. Dunleavy, Gideon Meyerowitz-Katz, Alberto Antonietti, Jose A. Calvache, Mark J. Kelson, Meg G. Salvia, Camila Olarte Parra, Saman Khalatbari-Soltani, Taylor McLinden, Arthur Chatton, Jessie Seiler, Andreea Steriu, Talal S. Alshihayb, Sarah E. Twardowski, Julia Dabravolskaj, Eric Au, Rachel A. Hoopsick, Shashank Suresh, Nicholas Judd, Sebastián Peña, Cathrine Axfors, Palwasha Khan, Ariadne E. Rivera Aguirre, Nnaemeka U. Odo, Ian Schmid, Matthew P. Fox

## Abstract

We estimated the degree to which language used in the high profile medical/public health/epidemiology literature implied causality using language linking exposures to outcomes and action recommendations; examined disconnects between language and recommendations; identified the most common linking phrases; and estimated how strongly linking phrases imply causality.

We searched and screened for 1,170 articles from 18 high-profile journals (65 per journal) published from 2010-2019. Based on written framing and systematic guidance, three reviewers rated the degree of causality implied in abstracts and full text for exposure/outcome linking language and action recommendations.

Reviewers rated the causal implication of exposure/outcome linking language as None (no causal implication) in 13.8%, Weak 34.2%, Moderate 33.2%, and Strong 18.7% of abstracts. The implied causality of action recommendations was higher than the implied causality of linking sentences for 44.5% or commensurate for 40.3% of articles. The most common linking word in abstracts was “associate” (45.7%). Reviewer’s ratings of linking word roots were highly heterogeneous; over half of reviewers rated “association” as having at least some causal implication.

This research undercuts the assumption that avoiding “causal” words leads to clarity of interpretation in medical research.

## Introduction

Health sciences research often investigates the relationship between a particular exposure and an outcome. Causal effects between these variables are often implicitly of interest, including studies based on non-random assignment of the exposure. Most researchers are aware that inferring causality may be fraught with difficulty, and that cautious interpretation may be warranted. However, this “caution” often manifests itself as avoiding causal language, potentially at the expense of clarity regarding study objectives and the plausibility of the underlying causal assumptions.. Some author guidelines (e.g., Journal of the American Medical Association^1^) explicitly prohibit the use of causal language in any study that is not a randomized controlled trial (RCT), often justified by the inaccurate, but common, belief that causal inference is only possible with RCTs.^2,3^ Health scientists and editors often employ euphemisms or language workarounds.^4,5^ For example, researchers may reserve use of causal language for only some parts of the manuscript^6^ or use language that can pass as either causal or non-causal. Alternatively, non-causal language may be used throughout the manuscript, but practical recommendations may still be offered that suggest or require a causal interpretation.^7^ It is not entirely clear what “counts” as causal language, with no clear standards and few attempts^6,8–12^ to define and categorize what constitutes causal language.

The use of ambiguous language leads to potential disconnects between the authors’ intentions, methods, conclusions, and perceptions of the work by research consumers and decision-makers.^4,5,13,14^ It may also indirectly erode research quality by enabling researchers to make ambiguously causal implications without being accountable to the methodological rigor required for causal inference. Otherwise non-causal language may morph into causal language in outlets for medical practitioners,^7,10^ press releases,^15–17^ and media reports.^18,19^ Ambiguous language may also imply greater support for any practical recommendations that require causal interpretation.^20^ While some loss of nuance may be attributed to press officers, journalists, and news recipients, too-strong language often starts from the study publications themselves.^18^ Most importantly, choice of language impacts research consumers’ and decision-makers’ perceptions,^13^ which in turn impacts health decisions.

Despite widespread discussions about causal language use,^4,5,21^ systematic evidence of its usage in practice is limited. In a review of 60 observational studies that were published in The BMJ, a fifth were judged to have inconsistencies in their use of causal language.^6^ Prevalence and use of causal language has been examined in studies concerning the overall medical literature,^6,18,22^ obesity,^11^ and orthopedics,^23^ noting that in the latter all uses of causal language in non-RCTs were assumed to be “misuse.” To date, there have been no large-scale systematic assessments of language used to link exposures and outcomes in the medical and epidemiological literature; existing efforts^6,8–12^ heavily focus on binary assessments of the language used (causal vs. non-causal).

This study systematically examined the linking language used in studies with a main exposure and outcome in the high-profile medical and epidemiological literature. Our objectives were to (i) identify the linking words and phrases used to describe relationships between exposures and outcomes, (ii) generate estimates of the strength of causality stated or implied by the linking phrases and sentences using a guided subjective assessment process, (iii) examine the prevalence of action recommendations that would require causal inference to have been made, and (iv) examine disconnects between causal implications in linking sentences and action implications.

## Methods

Our target sample consists of studies that quantified the relationship between a main exposure and an outcome in humans and were published in high-profile general health, medicine or epidemiology journals between 2010 and 2019. Years 2020-2021 were not included due to disproportionate focus on the coronavirus disease of 2019 (COVID-19). The study was pre-registered on the Open Science Framework (OSF): https://osf.io/jtdaz/. Changes made to the protocol after preregistration are documented and explained in Appendix 1.

### Search

Our search was structured in two steps: a preliminary search for appropriate journals and a secondary search for published papers within these journals.

#### Journal inclusion/exclusion criteria

The “top” journals in health, medicine, and epidemiology were determined by journal ranking from journals listed under Journal Citation Reports (JCR)^24^ categories for medicine and public health and SciMago’s category for Medicine. The top 200 journals from the SciMago Journal rank (SJR)^25^ and JCR’s impact factor rating for medical journals, and the top 200 highest impact factor rating journals for Public Health as extracted on May 26, 2020 were screened according to the following inclusion criteria: (1) mainly serves articles that are peer-reviewed, about health-specific topics, mainly publishing original, non-meta-analyses, review, orother secondary research designs), mainly concerning human-level observations (e.g., not animal models or microbiology); (2) must be a general health/medicine/epidemiology journal (i.e., journals which are focused on a narrow speciality and/or disease area of medicine were excluded); (3) the journal must have been founded in 2010 or earlier.

Among the journals meeting these criteria, lists of the 15 highest ranked journals by (1) impact factor, (2) h-index, and (3) SJR score were combined into a single list without duplicates. An additional decision was made during screening on June 24, 2021 to drop journals that where fewer than <10% of articles screened met the inclusion criteria and/or that did not have sufficient remaining unscreened articles to meet the minimum quota of articles from a single journal (See Appendix 1).

#### Search terms

We searched PubMed to identify all articles published in an eligible journal between 2010 and 2019 inclusive (details in Appendix 3). Medical Subject Headings (MeSH) terms were used to eliminate articles not meeting inclusion criteria. The search was performed in R^26^ using the EasyPubMed package^27^.

Articles were stratified by journal and whether they had the “Randomized Controlled Trial” MeSH tag. Identified articles were sorted in journal/article type stratified random order for screening. Disease areas were obtained for each article using the 2020 MeSH tag hierarchy^28^ for disease area headings.

### Screening

#### Study inclusion/exclusion criteria

Studies were eligible for inclusion if they were mainly concerned with the quantitative association of a main exposure/outcome pair, as assessed by reviewers as below:

- Observations must be human- or at an aggregate group of humans-level of observation
  - The main research question must be to examine the causal and/or non-causal association between one main exposure concept and one main outcome concept
  - One main exposure/outcome can include multiple measures of the same or similar broad exposure and/or outcome concept.
    - Articles can include many exposures/outcomes, but focus in particular on one exposure/outcome pair as their main association of interest (e.g., in the title, in the study aims)
    - Articles that are about more than one main concept (e.g., searching for what ‘risk factors’ are associated with the outcome) were excluded.
- The main research question must be examined quantitatively
- The main study design must not be a review or meta-analysis, or other secondary study design.

Studies investigating more than one exposure/outcome set were excluded because (1) it would not be possible to assess a main exposure/outcome pair per study; (2) study objectives and designs could not easily be compared with other papers; and (3) it would impose additional strain on the management of the data and review.

#### Procedures

Articles were screened continuously for each journal until journal quotas were met with the addition of a small buffer used for training purposes and for replacement of articles rejected during review. The journal quotas were 65 non-RCT articles and 6 RCT articles per journal, totalling 1,278 articles (1,170 non-RCTs and 108 RCTs). The sample size was informed by informal explorations of sample datasets balanced against reviewer capacity. We did not perform a formal sample size calculation because: 1) this descriptive study does not involve substantial hypothesis testing, 2) the variance in the language to be analysed in this study is unknown and is one of the key objectives of this study, and 3) the larger the sample size, the more in-depth we can explore less frequently used language, so we aimed to fully exhaust the available review capacity.

Articles were randomly assigned to three of 18 screening reviewers, with two independent reviewers and one arbitrating reviewer. During screening, the arbitrating reviewer made the inclusion/exclusion decision only in cases where the two independent reviewers disagreed.

Screening reviewers were presented with a list consisting exclusively of titles and abstracts, without additional information. The order of the lists to review was sorted randomly, stratified by journal and study design type (i.e., RCT vs non-RCT). An administrator periodically consolidated completed screening reviews and assigned articles for arbitration when disagreements occurred. Once quotas for each journal were met, further screening of articles from those journals was automatically disabled. Reviewers at no time had knowledge of from what journal a given article to screen came.

### Main review

#### Reviewer recruitment and selection

Reviewers were recruited through a combination of personal and Twitter solicitations. Reviewers were selected from those who expressed interest based on relevant graduate school education, expertise in relevant areas (e.g., epidemiology, causal inference, medicine, econometrics, meta-science, etc.), availability, and to maximize the diversity of fields, life experiences, backgrounds, and kinds of contributions to the group. All reviewers who completed their assigned reviews are coauthors.

The plurality (n=16/48) of reviewers were doctoral level students, followed by postdoctoral fellows (n=12/48) and faculty (n=10/48). The majority listed epidemiology as one of their primary fields (n=27/48), followed by statistics/biostatistics (n=9/48), medicine (n=6/48), economics (n=4/48), psychology (n=4/48), among other fields. Twelve reviewers have formal clinical training, while 6 were currently practicing clinicians. A plurality of reviewers were based in the United States (n=18/48) followed by the United Kingdom (n=9/48), Germany (n=4/48), Australia (n=4/48), and Canada (n=4/48), among other countries. Additional details are available in Appendix 2.

#### Reviewer roles and training

All reviewers received one hour of instruction and an additional set of training articles to review before the independent review. During the training and main review periods, reviewers were encouraged to engage in an active discussion on Slack to clarify guidelines, discuss issues, and generate community standards for review areas that may be more ambiguous. Reviewers were instructed to avoid referring to specifics of a particular study and to instead keep the discussion in general terms to balance eliciting individual subjective opinions with group guidance. By design, reviewers may have changed their understanding of the guidance over time through discussion, and were therefore allowed to make changes at any point before arbitration.

Each article was reviewed by three unique randomly selected reviewers; two independent independent reviewers and an arbitrating reviewer. The arbitrating reviewer was given the submitted data from the independent reviewers. Rather than simply resolving conflicts, the arbitrating reviewer’s task was to generate what they believed to be the best and most accurate review of each article, given the information from both independent reviewers, their own reading, and the ongoing community discussions. Arbitrating reviewers were free to decide in favor of one reviewer over another, consolidate and combine reviewer responses, or overturn both independent reviewers as they deemed appropriate. The arbitrator review data represents the main output of the review process and was used for all subsequent analyses.

#### Review framework and tool

The review framework and tool were designed to elicit well-guided, replicable, subjective assessments of the key questions for our study. The framing and definitions of words used (e.g., definitions and guidance for how/why language might be “causal”) are provided in Appendix 4.

Reviewers had the option to recuse themselves of reviewing each article for any reason (e.g., conflicts of interest, connections to authors, etc.); the article was then reassigned to another reviewer. Reviewers could also request that an administrator reevaluate the inclusion of a study. If the administrator determined that the article did not meet inclusion criteria, it was replaced with one from the buffer of accepted screened reviews.

Reviewers first identified the main outcome and exposure, preferably from the title of the study. Reviewers were asked to identify and copy and paste the main linking sentence, which generally was a sentence in the conclusions section of the abstract or full text containing the primary exposure, outcome, and the linking word/phrase. A linking word/phrase is defined as a word or phrase describes the nature of the connection between some defined exposure and some defined outcome as identified by the study analysis. This can describe the type of relationship (e.g., “associated with”) and/or differences in levels (e.g., “had higher”) that may or may not be causal in nature. Then, reviewers were asked to identify modifying phrases, or any words/phrases that modify the nature of the relationship in the linking phrase. This includes signals of direction, strength, doubt, negation, and statistical properties of the relationship (e.g., “may be”, “positively”, “statistically significant”).

Reviewers assessed the degree to which the linking sentence implied a causal relationship between the exposure and outcome using a four point scale (“linking sentence causal strength”) shown in Table 1.

**Table 1:** Causal implication strength rating scale

| Rating | Linking sentence | Action recommendation |
| --- | --- | --- |
| N/A | - | No action recommendation exists. |
| None | The linking sentence does not imply in any way that a causal relationship was identified. | The action recommendation would be made appropriately in the absence of any causal relationship. |
| Weak | The linking sentence might imply that a causal relationship was identified, but it is unclear or possible to come to that conclusion in the absence of any causal inference. | The action recommendation may be made appropriately had a causal relationship been identified, but it is unclear or possible to come to that recommendation in the absence of any causal inference. |
| Moderate | The linking sentence mostly implies that a causal relationship was identified, but it is unclear or possible to come to that conclusion in the absence of any causal inference. | The action recommendation most likely could only be made appropriately had a causal relationship been identified, but it is unclear or possible to come to that recommendation in the absence of any causal inference. |
| Strong | The linking sentence clearly implies that causality had been identified. | The action recommendation could only be made appropriately had a causal relationship been identified. |

Next, reviewers were asked to identify any sentences that contained action recommendations (how a consumer of the research might utilize the results and conclusions of the research). This may include recommending that some actor(s) consider changes (or no changes) in some set of procedures and actions. General calls for additional research were not considered action recommendations. After identifying this sentence (if applicable), reviewers were asked to consider the extent that this recommendation would require that a causal relationship had been identified, shown in Table 1.

In this framing “no causal implication” does not imply “no or null effects.” Reviewers were instructed to consider causal implications conceptually separately from the size (or lack thereof) of associations and correlations. Strong causal implications may be made even if the effect size measured was null, so long as the language implied that a causal relationship was being examined.

All articles received a review of the titles and abstracts. In addition, one-third of the articles underwent full text assessment. This extended review 1) repeated the abstract review questions for the discussion section and any pop-out sections (i.e., sections that do not appear as part of the main text or abstract, but summarize and highlight key aspects of the study), and 2) included additional questions to help indicate potential areas of causal intent,^29^ (see supplementary data). Reviewers also extracted whether there was any theoretical discussion about causal relationships between the exposure and outcome in the introduction, the number of covariates controlled or adjusted for, whether confounding was explicitly mentioned by name,^14^ whether a formal causal model was used, and whether explicit causal disclaimer statements were made (e.g., “causation cannot be inferred from observational studies, but…”).

### Root linking words/phrases language strength

After arbitrator reviews were completed, we compiled and curated a list of words from the linking words/phrases in the arbitrator reviews, and manually stemmed into their root words. Reviewers then rated the causal implications of all root words that were found more than once in our sample. This was to mimic language decision processes that base their causal language assessment on selecting words that are or are not causal, and to establish our own systematic assessments of word ratings. Reviewers were presented with up to four randomly selected linking words/phrases that contained the root word and had been submitted by arbitrating reviewers (e.g., the root word “associate” had four phrases, including phrases like “associated with” or “association”).

### Analysis

The statistical analysis was largely descriptive (e.g., describing the distributions of key extracted variables). Except for comparisons between RCTs and non-RCTs, all statistical analysis was performed on the arbitrated dataset of the non-RCTs only.

Comparisons between two ordinal categorical variables (e.g., strength ratings for causal implications of linking sentence vs. action implications) were estimated by Spearman’s correlation coefficients. Associations between strength ratings and key binary variables (e.g., study type, journals, topic areas, etc.) were estimated with ordinal logistic regression.

All measures of statistical uncertainty were clustered by journal and calculated using a block bootstrapping procedure unless otherwise specified, where 95% confidence intervals (CIs) were obtained from percentiles of the bootstrapped estimate distribution. Where the journals themselves were covariates, the clustered sandwich estimator was used. For root word rating proportions, there were no journal clusters, and as such the Wilson estimator was used. No weights were applied (i.e., journals and articles respectively contribute equally to our main results).

Heterogeneity between reviewers was evaluated using Krippendorf’s alpha. For the purpose of this review, disagreement between reviewers is a key result (i.e., heterogeneity between subjective opinions), rather than error.

All data management and analyses were conducted using R v4.0.5.^26^ Spearman correlation coefficients were determined using the pspearman package.^30^ Ordinal logistic regression was performed using the MASS package.^31^

### Data and code availability

All data and code are publicly available through our OSF repository: https://osf.io/jtdaz, except for files containing personal identifying information and/or personal API keys.

### Patient and Public Involvement statement

No patients or participants were involved with this research. All data were obtained from academic literature sources.

### Ethics approval

This research is not human subjects research, and as such no ethical approval was required. This research complies with the Declaration of Helsinki.

## Results

### Search and screening

Figure 1 summarizes the selection of journals and articles into the sample. Eighteen journals were identified meeting our search criteria: American Journal of Epidemiology, American Journal of Medicine, American Journal of Preventive Medicine, American Journal of Public Health, Annals of Internal Medicine, BioMed Central Medicine, British Medical Journal, Canadian Medical Association Journal, European Journal of Epidemiology, International Journal of Epidemiology, Journal of Internal Medicine, Journal of the American Medical Association, Journal of the American Medical Association Internal Medicine, The Lancet, Mayo Clinic Proceedings, New England Journal of Medicine, PLOS Medicine, and Social Science and Medicine.

**Figure 1:**
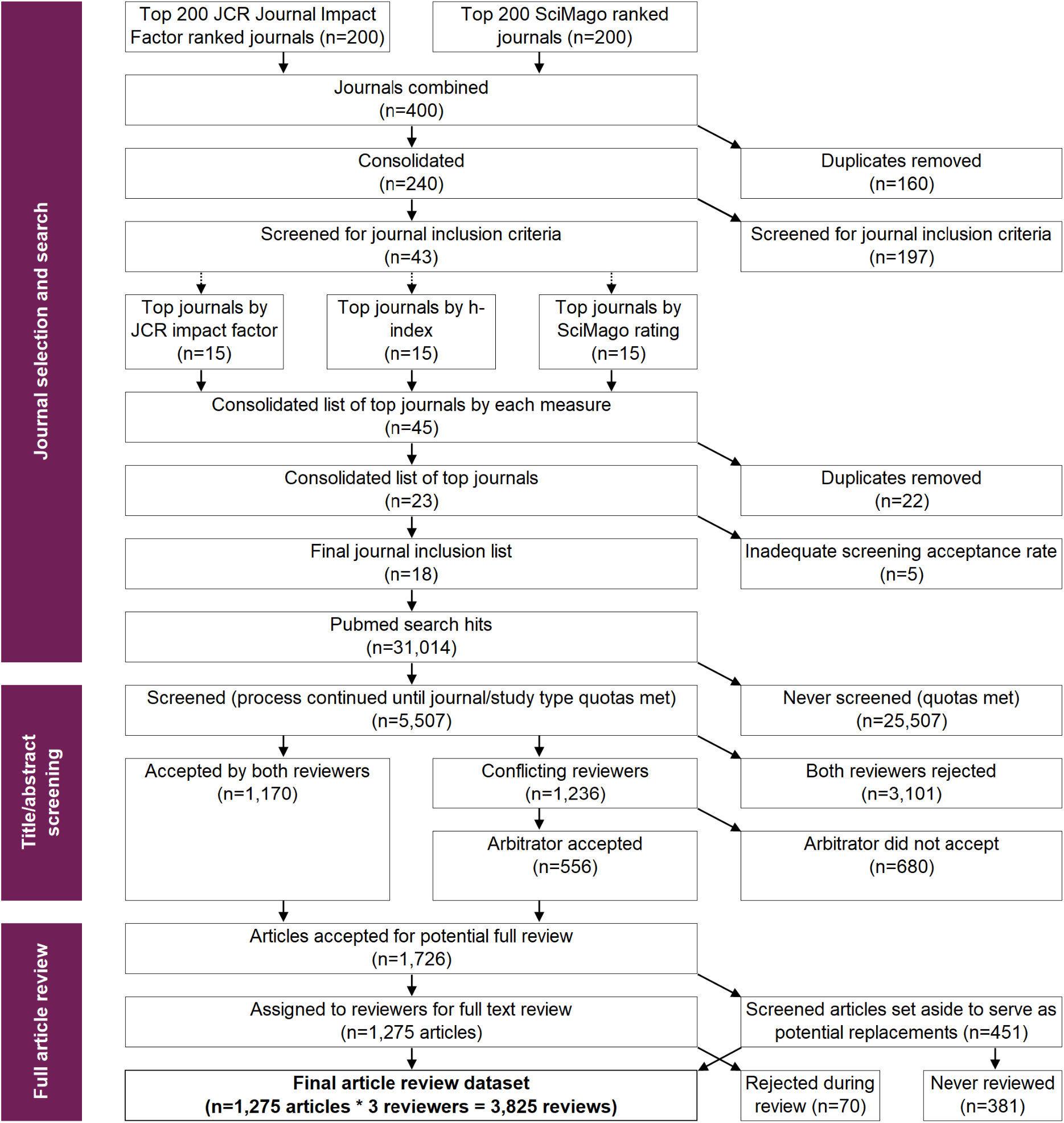
PRISMA diagram showing the selection of journals and articles into the study sample This chart shows the PRISMA diagram detailing the search and screening process to arrive at our final sample.

After searching PubMed for articles published in these journals from 2010-2019, we screened articles until 65 non-RCTs and 6 RCTs were accepted from each journal except the European Journal of Epidemiology, where only 3 RCTs were identified and included. This yielded 1,170 non-RCTs and 105 RCTs, totalling 1,275 studies reviewed. There were 10 recusals recorded during the main review. The three most common MeSH disease areas were “Pathological Conditions, Signs and Symptoms” (n=377), “Cardiovascular Diseases” (n=324), and “Nutritional and Metabolic diseases” (n=198). See Appendix 5 for full terms.

#### Linking words and phrases

After the arbitrator reviews were completed, root words were obtained through stemming the linking phrases to identify and rate the root linking words.

By far the most common root linking word identified in abstracts was “associate” (n=535/1,170; 45.7%, 95% CI 40.0, 51.9%), followed by “increase” (n=71/1,170; 6.1%, 95% CI 4.7, 7.8%) (Figure 2). The same root word was identified in both the abstract and discussion for 48.2% cases (95% CI 43.7, 53.6%). We found only 9 (0.8%, 95% CI 0.4, 1.3%) studies where the main root linking word was “cause.” There were 16 (1.4%, 95% CI 0.6, 2.3%) articles that used the word “cause,” when additionally including any instance of the word “cause” in either the linking or modifying phrases.

**Figure 2:**
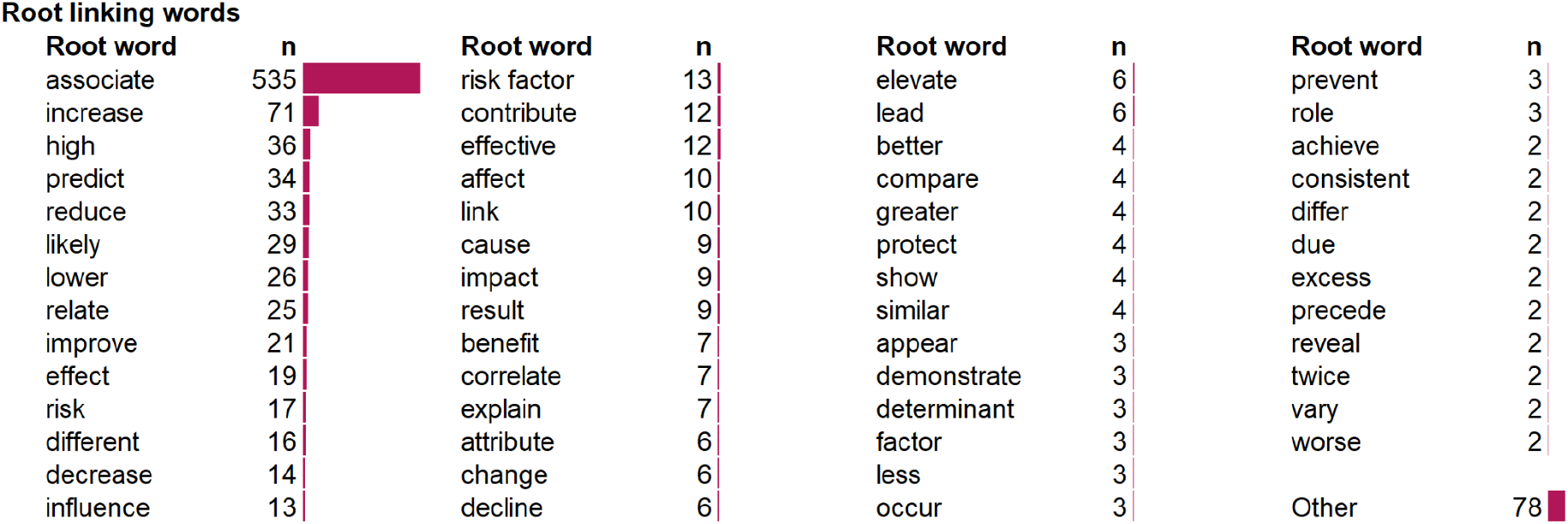
List and frequency of identified root words used to link the exposure and outcome This chart shows the number of times each of these root words appears in the linking phrases in the abstracts of our samples. In cases where two of these words are in the same phrase (e.g., “similar risk”) the more common of the two is selected (in this case “risk”). In cases where selected linking phrases had two or more words which were included in the root word list, the more common word was selected as the root word mainly associated with that study and section.

### Causal implication(s) strengths

#### Summary data

Reviewers rated the abstract linking sentence as having no causal implication in 13.8% (95% CI 11.9, 15.9%), weak in 34.2% (95% CI 31.4, 36.7%), moderate in 33.2% (95% CI 29.8, 36.7%), and strong in 18.7% (95% CI 15.1, 22.6%) of instances Figure 3). About half (n=8/18) of the journals had pop-out sections in their articles. The language used was very similar in the abstract, full-text discussion, and pop-out sections.

**Figure 3:**
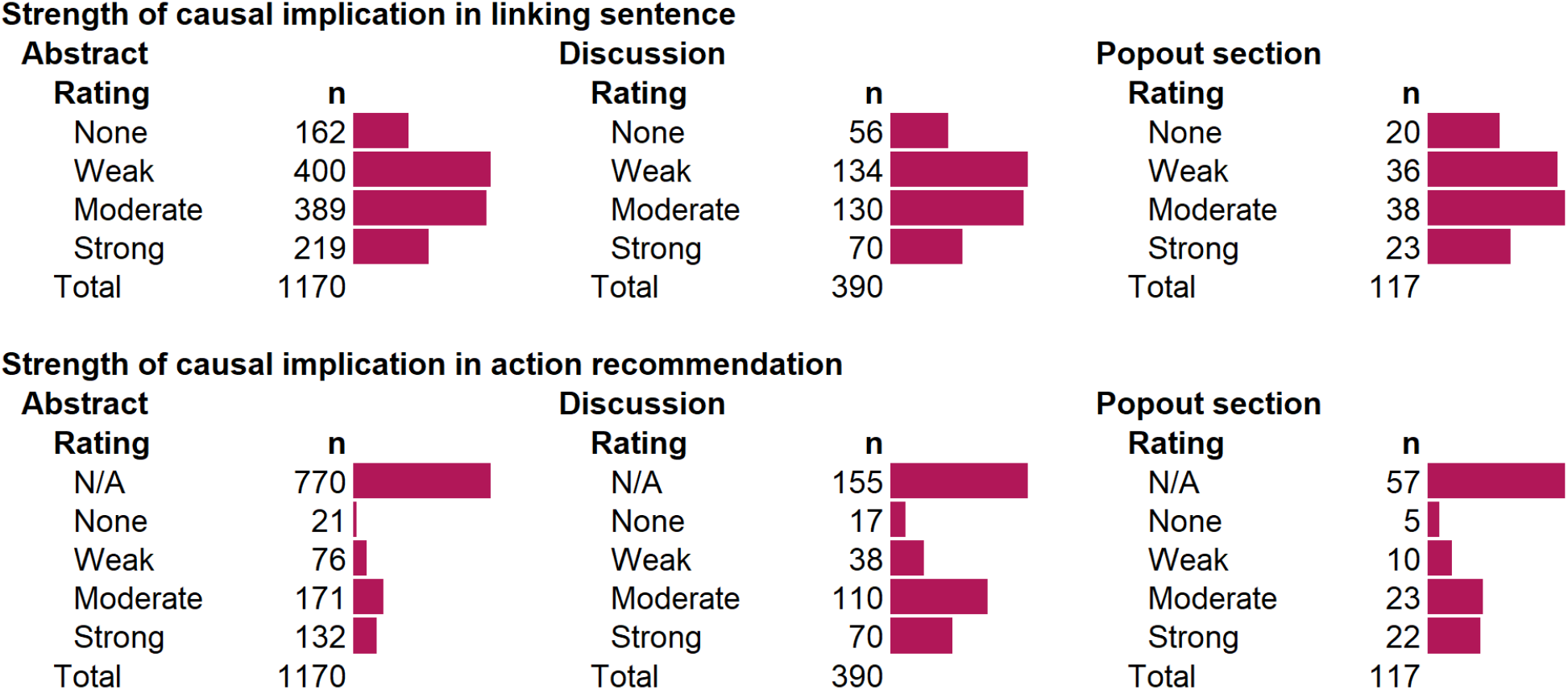
Summary scores for the degree of causal implication in linking sentences and action recommendations This chart shows the frequency of key strength of causal implication metrics for the 1,170 non-RCT studies in our sample, as indicated by the arbitrating reviewer.

Action recommendations were identified in 34.2% (95% CI 29.0, 39.6%) of abstracts. Of these, 5.3% (95% CI 3.5, 7.2%) were rated as having a causal implication of None, 19.0% (95% CI 15.2, 23.0%) Weak, 42.8% (95% CI 39.0, 46.4%) Moderate, and 33.0% (95% CI 29.0, 37.1%) Strong.

By comparison, action recommendations were identified in 60.3% (95% CI 52.7, 67.5%) of discussion sections, about twice that in abstracts. We found negligible, if any, differences between the overall strength of the action implications found in discussions sections vs abstracts [log odds for higher rank: -0.00026 (95% CI -0.00024, 0.00013)]. There was also no apparent pattern in implication strength over time (Appendix 6).

#### Comparison of linking sentence strength vs. action implication strength

Figure 4 shows the strength of causal implication for linking sentences and action recommendations among those studies with an action recommendation in the abstract (n=400, 34.0%). Of studies with action recommendations, 15.3% (95% CI 11.7, 19.2%) had action recommendations that less strongly implied causality than the linking sentence, 40.3% (95% CI 35.1, 45.8%) were commensurate, and 44.5% (95% CI 39.9, 48.4) were stronger (Panel A). There was a weak correlation between the strength of causal implication in the linking sentence and the action recommendation (Spearman’s correlation coefficient=0.349, 95% CI 0.256, 0.435)]. While stronger causal action recommendations are less likely to occur when linking sentences are weaker (Panel B), studies with weaker linking sentences also often make strong causal action implications. Among the 76.0% of studies with no action recommendation in the abstract, 14.5% (95% CI 11.6, 17.6%) of linking sentences were rated as having a causal implication of “None”, 34.0% (95% CI 30.3, 37.5%) Weak, 33.1% (95% CI 29.2, 37.3%) Moderate, and 18.3% (95% CI 14.5, 22.5%) Strong. We found negligible, if any, differences in the strength of the linking sentences between abstracts that did and did not contain action recommendations (log odds for higher rank: 0.087, 95% CI -0.162, 0.320).

**Figure 4:**
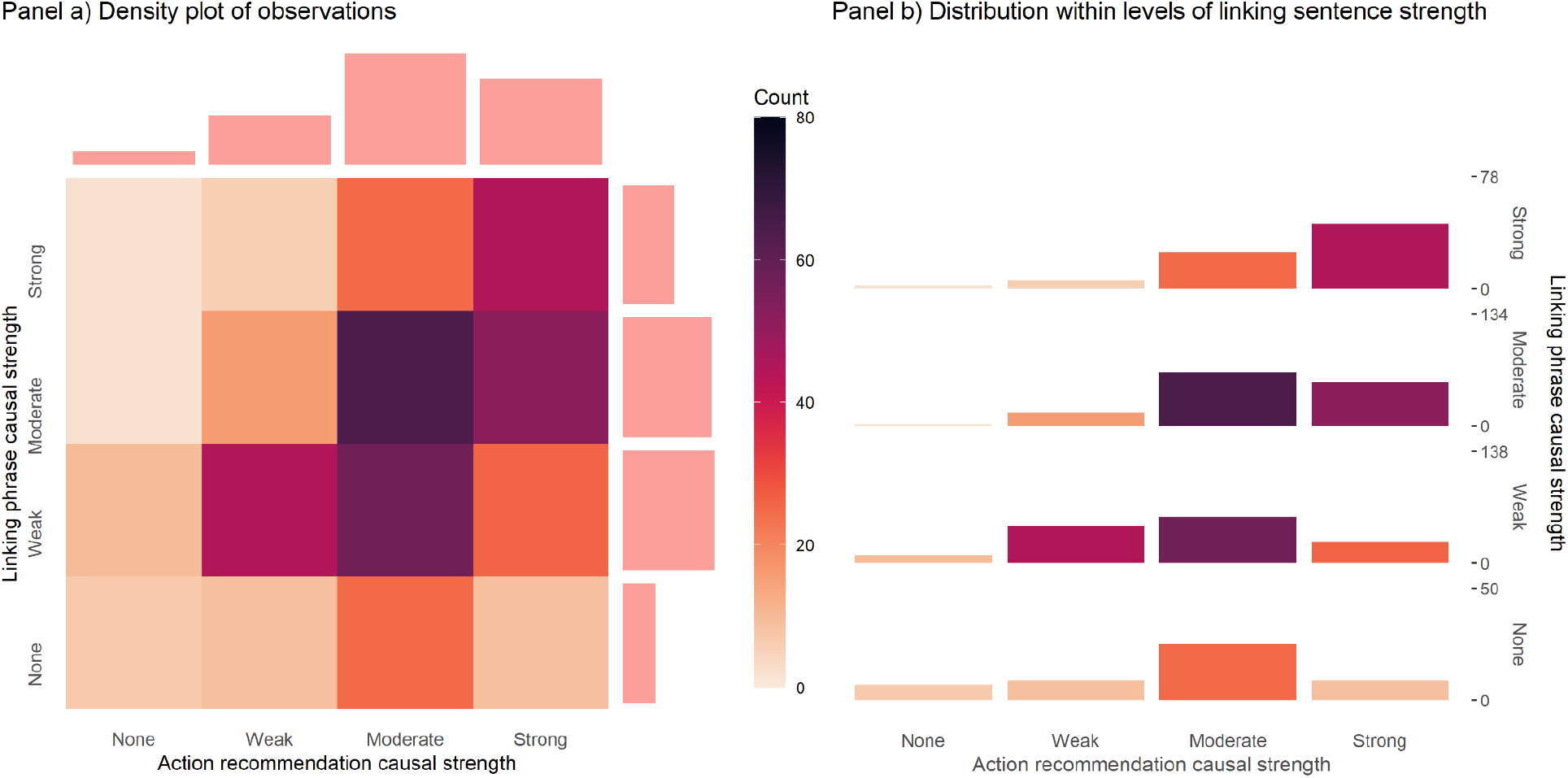
Comparison of the strength of causal implications in the abstracts for the linking phrase and action recommendation This chart shows the distribution of linking sentence and action recommendation language, among the 400/1,170 non-RCT studies in which there was an action recommendation present in the abstract. Panel A shows an unconditional heatmap, with colors representing the number of articles in the strata, and histograms on the top and right showing the overall distribution of ratings for each axis. Panel B shows the distributions within each level of linking sentence causal strength.

### Words and phrases

As shown in Figure 5, ratings among reviewers (n=47) for causal implication of root words were highly heterogeneous, with the only word to reach near consensus on causal implications being “cause” (Figure 5). Reviewers rated words such as “correlate” and “associate” generally rated weaker, in terms of their causal implications, than words such as “impact”, “effect”, “affect”, and “prevent.” Notably, many root words could be used in a variety of ways with potentially different meanings. For example, the root word “lower” could be used purely descriptively, as in “people with X had lower Y”, or indicating X as a driving force, as in “X lowered Y.”

**Figure 5:**
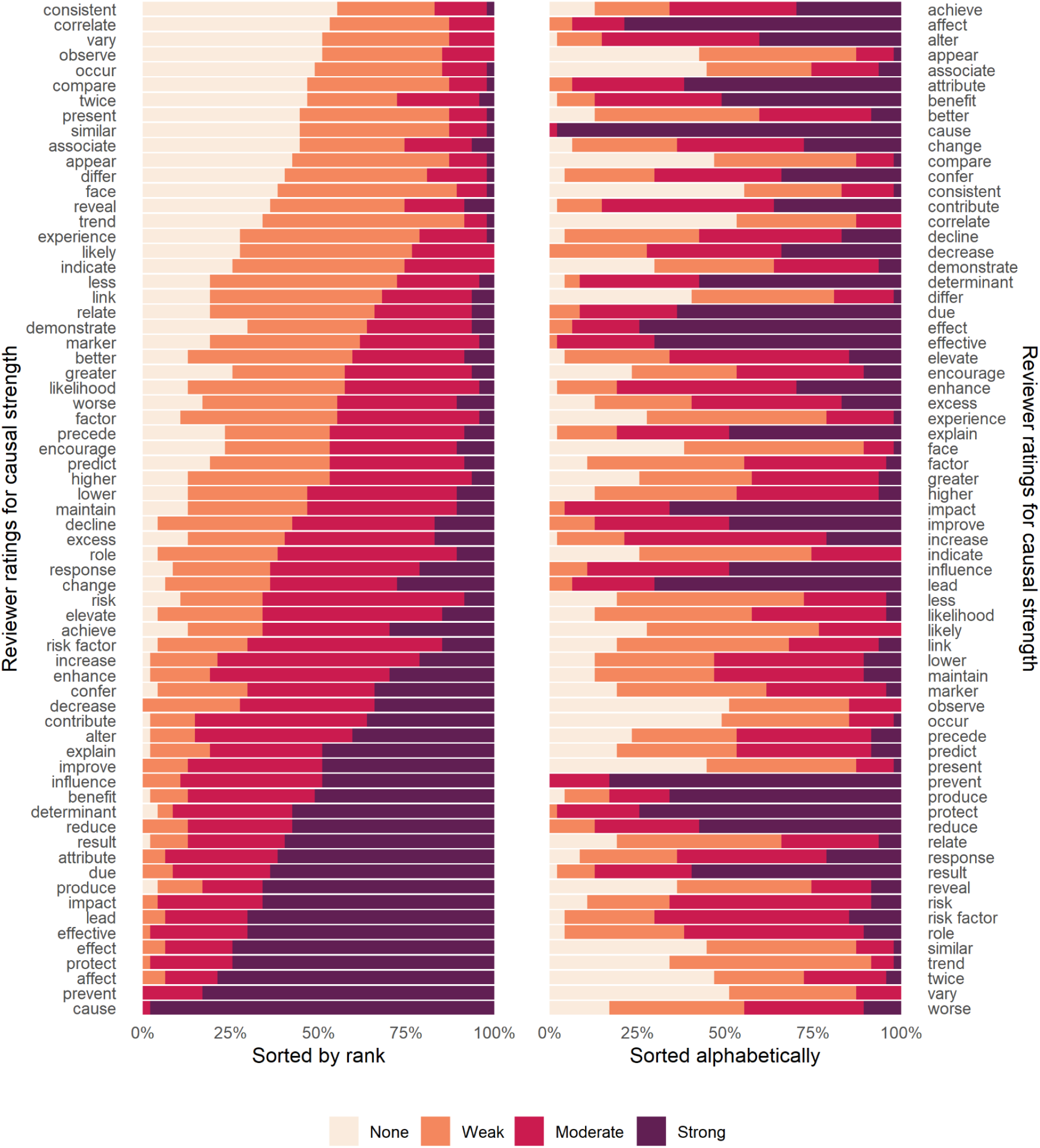
Strength of causal implication ratings for the most common root linking words This chart shows the distribution of ratings given by reviewers during the root word rating exercise. On the left side, they are sorted by median rating + the number of reviewers who would have to change their ratings in order for the rating to change. On the right, the chart is sorted alphabetically.

Although the word “associate” ranked among having the lowest overall causal implications, more than half of the reviewers judged that the word “associate” carried at least some causal implication [n=26/47, 55.3% (95% CI 41.2, 68.6%). For comparison, 78.6% (95% CI 75.7, 81.2%) of linking sentences containing the root word “associate” were rated as having at least some causal strength.

#### Modifying words and phrases

Modifying phrases were identified in the abstracts of 72.1% of studies (95% CI 69.0, 75.6%). 11.2% (95% CI 8.6, 14.2%) of studies had a modifying phrase with variations on “statistical” and/or “significant.” Phrases expressing caution (e.g., “may be,” “could,” “potentially”) or strength (e.g. “strongly,” “substantially,”) were both fairly common in the modifying phrases extracted. However, given the wide variety of phrases extracted and the lack of a pre-established framework for doing so, no formal categorization of modifying phrases was performed or quantified. The frequency of modifying words and phrases identified three or more times are shown in Appendix 7.

### Differences in strength across key strata

#### Non-RCTs vs. RCTs

For the RCTs, reviewers rated the causal implication of the abstract linking sentence as being None for 9.5% (95% CI 4.8, 15.2%), Weak for 6.7% (95% CI 2.8, 11.4%), Moderate for 27.6% (95% CI 19.0, 36.4%), and Strong for 56.2% (95% CI 46.4, 65.8%). This is overall much stronger than for the non-RCTs [log-odds of RCTs having higher rank: 1.63 (95% CI 1.26, 2.04)].

Overall, 75.2% (n=79/105; 95% CI 66.7, 82.9%) of RCTs in our sample had no action recommendation. Of the 26 that did, 0 were rated as having a causal implication of None, 6.7% (95% CI 2.8, 11.4%) Weak, 27.6% (95% CI 19.4, 36.6%) Moderate, and 56.0% Strong (95% CI 45.7, 64.8%). The action recommendation causal implications in the RCTs appeared stronger than in the non-RCTs [log odds for higher rank: -0.398 (95% CI -0.916, 0.009), noting that this is underpowered due to insufficient RCTs with action recommendations to make reasonable inference about differences.

The most common linking word identified in RCT abstracts was “associate” (n=16/105), followed by “reduce” (n=14/105), and “increase” (n=11/105).

#### Journals and journal policies

The strength of causal linking language and action recommendations was generally similar between journals (Appendix 8). Three journals have publicly posted policies regarding causal language. The Journal of the American Medical Association (JAMA) and JAMA Internal Medicine explicitly restrict the use of “causal” language to RCTs, while the American Journal of Epidemiology (AJE) discourages the use of the word “effect” exclusively, giving guidance as to when it should be used. Compared with the other 15 journals, the strength of the causal language in these three journals was weaker, especially for JAMA which had the lowest rank of any journal [log odds of having higher rank: -0.627 (95% CI -0.771, -0.483) for JAMA, -0.083 (95% CI -0.235, 0.069) for JAMA Internal Medicine, and -0.080 (95% CI -0.229, 0.069) for AJE. The difference in the causal language strength in named journals of epidemiology compared to other journals appears to be small, with the log odds of having a higher linking language strength being -0.140 (95% CI -0.447, 0.166)].

There was little difference between journals in causal strength of action recommendations when present, but there were some differences in their prevalence (Appendix 9 and Appendix 10).

Papers in JAMA, JAMA Internal Medicine, and AJE were less likely to include actions recommendations than papers in the other 15 journals [log odds for having action recommendations: -0.624 (95% CI -0.885, -0.363 for JAMA, -0.090 (95% CI -0.351, 0.170) for JAMA Internal Medicine, and -0.806 (95% CI -1.067, -0.546) for AJE]. Articles from epidemiology journals (n=3) were less likely to include action recommendations than the remaining (n=15) journals [log odds for having action recommendations -0.516 (95% CI -0.870, -0.163)].

### Indications of potential causal interest

Most studies in our sample provided *some* indication of potential causal interest (Figure 6). While only 3.8% (95% CI 2.0, 6.0%) of studies presented formal causal models, most offered some discussion of the theoretical nature of the causal relationship between exposure and outcome (80.0%; CI 75.2, 85.4%). Among those that discussed theory, 58.7% (95% CI 51.4, 64.8%) moderately or strongly indicated a theoretical causal relationship between the two. 24.6% (95% CI 20.9, 28.0%) of studies had a disclaimer statement regarding causality. 68.7% (95% CI 63.3, 73.7%) mentioned “confounding” explicitly (i.e. using the word “confound” or variations of it). Finally, most studies in our sample controlled or adjusted for several variables, with 35.1% (95% CI 30.5, 39.9%) having 10 or more control variables.

**Figure 6:**
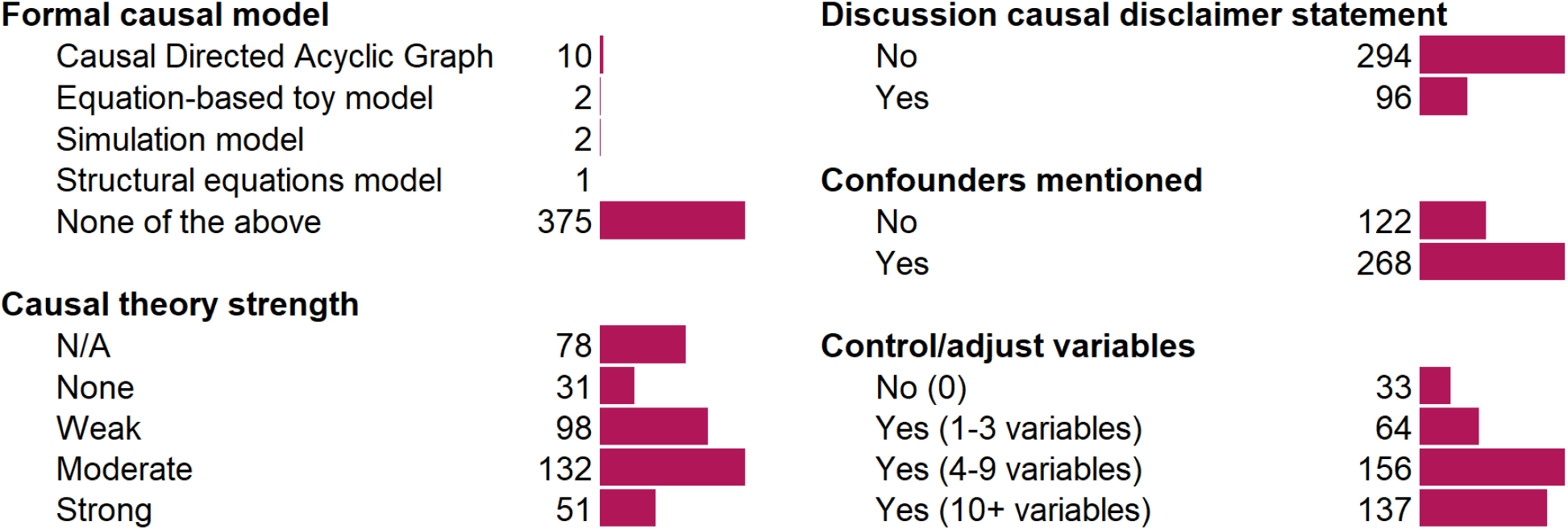
Frequency of indicators of potential causal interest These results are from the 390 articles reviewed in full.

### Inter-rater comparisons

The Krippendorff’s alpha comparing the reviewers’ ratings for linking language strength in the abstract was 0.29. independent reviewers gave the same score in 35.1% of instances; 41.2% differed by one category, 19.9% by two categories, and 3.8% by three categories. Agreement increased to 0.41 when including the independentand arbitrating reviewers.

For the action recommendations (where most articles were rated as “N/A” for missing) the Krippendorff’s alpha was 0.70. The two independent reviewers agreed exactly in 67.6% of cases, differed by one category in 14.4% of cases, by two in 8.6%, by three in 5.3%, and by four in 4.1%. Similarly, agreement increased to 0.76 when including the arbitrating reviewers.

## Discussion

Our systematic evaluation of the use of causal language and implications in the high-profile medical and epidemiological literature found that 1) by far the most common word used linking exposures and outcomes was “associate,” 2) although few studies explicitly declared an interest in estimating causal effects, the majority used language that moderately or strongly implied causality, 3) while only about a third of articles issued action recommendations, the vast majority of these were found to imply that causality had been inferred, and 4) causal language in action recommendations ratings tended to be stronger than the language in linking sentences, and 5) Although many studies used disclaimers warning readers against making causal inferences, an implicit interest in causality apparent from common discussions of causal mechanisms and widespread adjustment for confounding. Overall, we found a substantial disconnect between the causal implications used in technical linking language and research implications.

Our results suggest that “Schrödinger’s causal inference,” ^32^ - where studies avoid stating (or even explicitly deny) an interest in estimating causal effects yet are otherwise embedded with causal intent, inference, implications, and recommendations - is common in the observational health literature. While the relative paucity of explicit action recommendations might be seen as appropriate caution, it also invites causal inference since there are often no useful and/or obvious alternative (non-causal) interpretations. To our surprise, we found that the RCTs in our sample used similar linking words to the non-RCTs. Our review suggests that the degree of causal interpretation for common linking words has been impacted by the unavailability of explicitly causal language, such that the meaning of traditionally non-causal words has broadened to include potentially stronger causal interpretations.^33^ It is likely that the rhetorical standard of “just say association” has meant that many researchers no longer fully believe that the word “association” just means association.

At this time, we do not know the degree to which journal editors, reviewers, authors, or academic community standards contribute to the implicit and explicit rules of causal language. While there are relatively few explicit and public rules governing language at journals, journals may employ formal internal guidelines and unspoken informal norms.

Our measures of causal implication are based on subjective assessments, which are critical to evaluating and interpreting human language. Reviewers substantially differed regarding the causal implications of many linking words, even in the presence of extensive guidance, processes, and training for how to assess causal implication in language. Different interpretations may arise from different backgrounds, experiences, and other factors affecting personal interpretations. Our reviewers, for example, are likely to have selected into our study due to an interest in and knowledge of causal inference and language. Outside of this study, we expect that alternative potential target populations of consumers of health research (clinicians, policy-makers, and others) may also interpret these words differently, whether by virtue of differing frameworks for assessing language, personal interpretations, or community standards. Rather than attempting to be fully representative of any one possible population of people who interact with this research, we chose to have a co-author reviewer pool with representation from a wide variety of possible target populations. Our reviewer pool is not representative of any particular population, but instead covers a wide variety of research traditions that might interact with this type of research.

Beyond the reviewers themselves, it also matters how words are used and in what context. For example, we found that ratings between “associate” alone in the root word rating exercise had less causal implication compared with in-context ratings of sentences with “associate” in the linking phrase. Aspects of the rating and interpretation process are also likely to be particularly challenging; for example, in reviewer discussions many reported difficulty with evaluating the degree of causal implication for sentences with null findings. Research consumers and decision-makers may have entirely different interpretations and frameworks, consciously or otherwise.

This study was designed with replicability in mind. The review process was designed to balance independent subjective assessments from skilled researchers and practitioners with explicit guidance and discussion among reviewers. Our assessment process is applicable to any number of areas of systematic evidence review and evaluation, which is often limited to shallow “objective” measures. Beyond pre-registration, nearly all parts of this project were fully open and disseminated to the public to view and comment, including documents, data, and code, resulting in a very large number of contributors, comments, and suggestions throughout the process.

Results may not be directly generalizable to other settings, alternative samples, and reviewers. Because our inclusion criteria excluded studies that were examining several potential factors or exposures and their relationships with outcome(s), our sample likely excluded many multi-exposure articles with terms such as “risk factors,” “correlates,” or “predictors.” Our journal selection also included only the most prominent general medical, public health, and epidemiology journals, and may not be representative of different fields, subfields, journals and policies. We did not examine the strength of evidence, nor did we examine any information that would indicate the appropriateness of claims.

The practice of avoiding causal language linking exposures and outcomes appears to add little if any clarity. Common standards for which words and language are “causal” or when “causal” words are appropriate do not appear to match interpretation. While being careful about what we claim is critical for medical science, being “careful” is often implemented by stripping out causal language in conclusions, and therefore any hint of what question is being answered. Knowing that the association between X and Y is 42 is not informative if we do not know what question that association attempts to answer.^34^ Misalignment between the research question being asked and action implications is on its own a source of confusion, which could be avoided if the causal nature of the research question was made explicit. Further, these practices may weaken methodological accountability, as studies that only indirectly imply causality can be shielded from critiques regarding lack of causal inference rigor.^4^

Rather than policing which words we use to describe relationships between exposures and outcomes, we recommend focusing on how researchers, research consumers, and reviewers can better identify and assess causal inference designs and assumptions. Quantitative empirical research should clearly state its target estimand to clarify the research question (Tennant et al., 2021), including explicitly stating when such estimands are causal. Authors, reviewers and editors should focus on being clear about what questions are being asked,^35,36^ what decisions are being informed, and the degree to which we are and are not able to achieve those goals.

## Acknowledgements

This work was supported by many people who made contributions to this work. Turki Althunian contributed to the screening process. Jess Rohmann contributed to the piloting process. This work was additionally supported by comments and contributions from Alyssa Bilinksi, Pascal Goldsetzer, Caroline Blaine, Otto Kalliokoski, Eero Raittio, Tanya Colyer, Tim Watkins, Alexander Breskin, Arindam Basu, Jessica L. Rohmann, Luke A McGuinness, Todd Johnson, Mario Malički, Sebastian Skejø, Scott Graham, Michael Chaiton-Murray, John Edlund, Katelyn Smalley, Danielle Newby, Anita Williams, Cord Phelps, Colleen Derkatch, Alexander Wolthon, Pallavi Rohella, Damien Croteau-Chonka, Steven Goodman, and John Ioannidis.

All errors are the sole responsibility of the authors.

## Author roles

Protocol design: NAH, SW, MPF, JMR, OAA, PWGT, EJM, EAS

Study administration: NAH, SW

Data management: NAH

Statistical analysis: NAH

Graphical design: NAH

Study design (piloting): SW, SPi, ER, CL, ALO, RB, SD, MDLRT, TSA, DJD, MS, TM, SPe Data analysis (screening): NAH, SPi, STL, SJH, AES, PS, AB, MSKD, SD, TRE, DRM, TMA, GMK, AA, JAC, MJK, COP

Data analysis (main review): NAH, SW, MPF, JMR, OAA, PWGT, EAS, SPi, STL, ER, SJH, AES, CL, PS, AB, MSKD, ALO, RB, SD, MDLRT, TRE, DRM, TMA, DJD, GMK, AA, JAC, MJK, MS, COP, TM, AC, JS, AS, TSA, SET, JD, EA, RAH, SKS, SS, NJ, SPe, CA, PK, AERA, NUO, IS

Manuscript writing: NAH, SW, JMR

Manuscript editing: NAH, SW, MPF, JMR, OAA, EJM, PWGT, EAS, SPi, STL, ER, SJH, AES, CL, PS, AB, MSKD, ALO, RB, SD, MDLRT, TRE, DRM, TMA, DJD, GMK, AA, JAC, MJK, MS, COP, TM, AC, JS, AS, TSA, SET, JD, EA, RAH, SKS, SS, NJ, SPe, CA, PK, AERA, NUO, IS

NAH serves as the primary guarantor of all aspects of the study and takes full responsibility for the work.

## Transparency statement

The lead author, Noah A. Haber, affirms that this manuscript is an honest, accurate, and transparent account of the study being reported; that no important aspects of the study have been omitted; and that any discrepancies from the study as planned (and, if relevant, registered) have been explained.

## Appendices

## Appendix 1

### Changes from pre-registered protocol

Major changes:

1. The main measure of linking language causal implication strength indirectly through the root words to direct reviewer ratings of the sentences themselves.
  a. In the original protocol, the main method of generating causal implication for the linking language was through the root word rating system, where those ratings would then be applied back to the studies from which they came. No question was asked regarding the causal implications of the linking sentence in context.
  b. During piloting, we added the question to the review tool which had reviewers directly assess the causal implications of the linking sentences as a whole in order to better rate and review the language in context.
  c. During the independent review, but before the arbitrator review phase, we changed our main linking language measure from the root word exercise to the direct ratings of the sentences themselves.
  d. This decision was made for three reasons
    i. This greatly simplified the estimation of the main results, negating the need to back-apply causal language from the root word ratings
    ii. The full sentence context would be a more direct and contextually sensitive assessment of causal language than the root word exercise.
    iii. During an interim data quality check of the reviewers’ extracted linking phrases, we found that the extracted data were much more heterogeneous than initially anticipated, lending some doubt whether the original strategy was viable and interpretable.
2. Journals with very low rates of screening acceptance were retroactively excluded from the list of journals
  a. This decision was made partway through the screening process itself.
  b. Because the protocol specified that we would have the same number of articles accepted from each of the journals, during the screening process we found that screeners would have to work vastly more to meet journal quotas among the journals which had very low rates of screening acceptance.
  c. On June 24, journals which had screening acceptance rates of below 10% or journals in which there were not enough unscreened articles remaining to meet quotas were excluded, and quotas were increased to compensate among the remaining journals.
  d. This decision was made for two main reasons:
    i. Keeping these journals would have created an infeasible amount of screening required to complete the screening process.
    ii. Journals with such low rates of screening acceptance were likely less relevant to meet our stated objectives and journal inclusion criteria.

Minor changes:

1. We selected 18 journals, rather than the initial expected 20 from the protocol.
  a. The expected number of journals in the protocol (20) was made in error. We chose to follow the process, rather than aim for a specific number of journals. This initially yielded 24 journals, 6 of which were later removed due to low screening acceptance rates (see above)
2. The sample size target changed to 1,170 non-RCTs (61 per journal) and 90 RCTs (6 per journal)
  a. The protocol was initially stated to be 1,525 articles accepted, with 61 non-RCTs per journal and 6 RCTs.
  b. This was reduced due to lower than expected screening acceptance rates in order to ensure that screening logistics were feasible and that schedules would be met.
3. The data extraction form received a large number of minor tweaks to the language, phrasing, and guidance.
  a. These changes were made as part of the protocol-specified piloting process.
4. The root word extraction process was performed on the linking phrases collected from the arbitrator reviews, rather than the independent reviews.
  a. This ensured a cleaner dataset of linking words and phrases from which to extract root linking words
5. Root words were only included in the root word linking exercise if there were two or more instances of them from the arbitrator reviews, and a light curating process was performed afterwards.
  a. This was performed due to clean up highly heterogeneous extracted linking words and phrases
6. The root word rating exercise was changed to being performed after the arbitrator reviews.
  a. In the original protocol, the root word exercise occurred during the arbitrator reviews.
  b. This change was made in order to accomodate extracting the root words from the arbitrator-extracted linking phrases
7. The reviewers were assigned to review all of the words in the root word list
  a. The original protocol specified that the reviewers would only review 20 randomly selected root words
  b. This was performed in order to maximize the power of our sample.
8. Spearman’s correlation coefficients were added to directly examine the correlation of rankings between ordinal categories
  a. This was not originally specified in the protocol due to oversight, and was added later.
9. The population weighted tertiary analysis was removed
  a. This was omitted due to lack of clear value of targeting an alternative “population” of studies, to simplify the breadth of analyses, and due to lack of space

## Appendix 2

### Reviewer characteristics chart

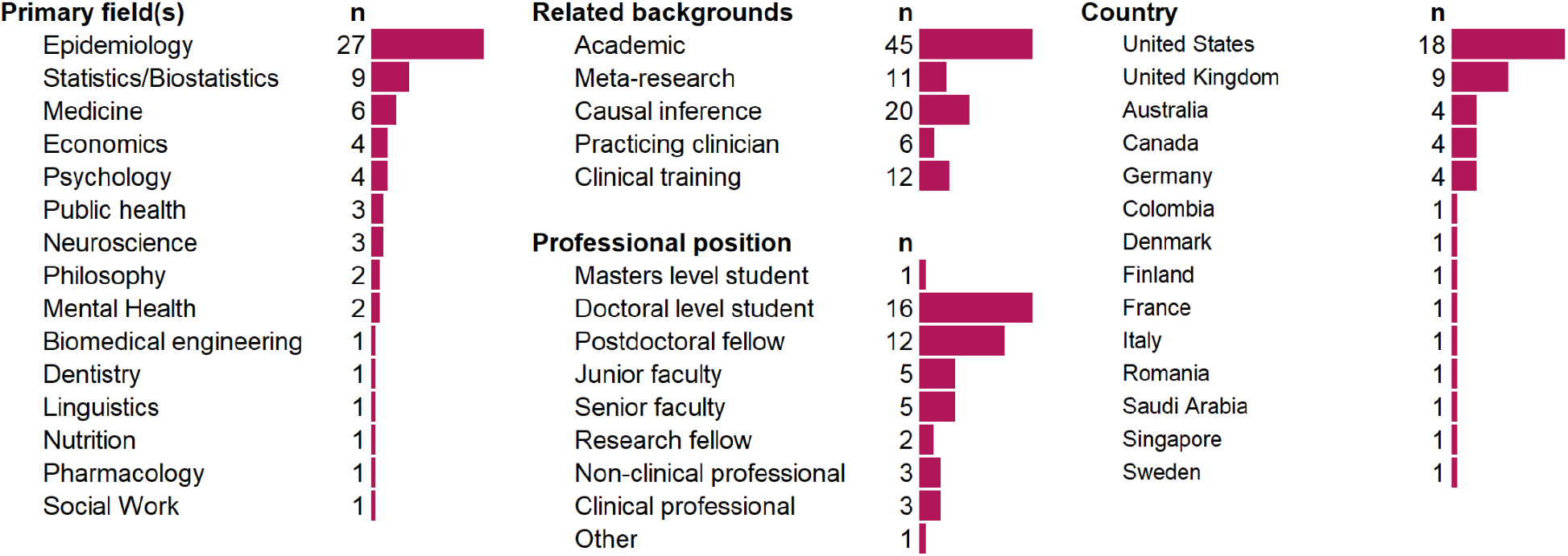

## Appendix 3

### Search terms

Our search was performed and pulled from PubMed to extract title, abstract, MeSH keywords, and citation data, using the following terms:

> ((*<year>*[PDAT]) AND (*<journal ISSN>*[Journal])
>
> AND
>
> (Humans[mesh] AND “Journal Article” [PT] AND English [la] AND hasabstract))
>
> NOT
>
> ((“Meta-Analysis”[Publication Type] OR “Review”[Publication Type] OR “Case Reports”[Publication Type] OR “Editorial” [Publication Type] OR “Letter”[Publication Type]))”

Where *<year>* is the years from 2010 to 2019, and *<journal ISSN>* is the journal in question. The above search was performed for every year/journal combination and combined.

## Appendix 4

### Definitions and frameworks

#### Exposure

For this project, “Exposure” refers to the independent variable of interest (in a regression sense) or the main or antecedent variable being investigated for a possible (non-)causal link to the study outcome, or resulting or end-point variable. It may be labelled by terms such as treatment, factor, risk factor, protective factor, determinant, intervention, correlate, predictor, agent, cause, causative agent, or other terms.

#### Outcome

For this project, “Outcome” refers to the dependent or effect variable of interest that is being investigated for a possible link to the exposure (surrogate measures or clinical events). It is typically a post-exposure variable i.e. assumed or known to be preceded by the exposure. It is sometimes called the study endpoint variable, consequence, result, or other terms.

#### Linking word/phrase

A linking word/phrase describes the nature of the connection between some defined exposure and some defined outcome, generally used in a sentence containing both exposure and outcome. This can describe the type of relationship (e.g. “associated with”) and/or differences in levels (e.g. “had higher”) that may or may not be causal in nature. For our purposes, the phrase may contain 1-3 words, where one of the words is a preposition to link the exposure and outcome. Some examples may include constructions such as “associated with,” “effect of,” “increased,” “was higher than,” “correlated with,” “caused,” “harms,” “predicts,” “risk factor for,” “determined,” “impacts,” “decreased,” “linked to,” etc.

#### Modifying word/phrase

A modifying word/phrase is a word or phrase that modifies the linking word/phrase describing the nature of the relationship between the exposure and outcome. This includes adding signals of direction, strength, doubt, negation, and statistical properties to the relationship. This may include phrases like “may be,” “positively,” “strongly”, “potentially”, “is likely to,” “does/is not,” “statistically significant,” etc.

#### Causal language

Causal language implies that one entity influences (or does not influence) another. We define language as being causal if that language implies that movement (or lack thereof) in the outcome was either 1) impelled by the exposure of interest (i.e. a change in the exposure drives or does not drive a change in the outcome, e.g., increase, decrease, improve, change), or 2) implies attribution of the outcome to the exposure (i.e. assigns the responsibility for the change or lack of change in the outcome to the exposure, e.g. “due to,” “since,” “attributable to”).

#### Action recommendation

This is a description of how a consumer of the research in question might utilize the results and conclusions of the research. This may include recommending that some actor consider changes (or no changes) in some set of procedures and actions. Action recommendations concern what to do with the research. For our purposes, we do not count calls for additional research as action recommendations.

#### Causal implication of recommendations

Recommendations may often imply a causal interpretation of a finding. For example, authors may suggest that it could be beneficial to change the amount of an exposure, which rests on the assumption that the exposure has a causal effect on the outcome. As a variation, it may also be suggested that an exposure need not be changed, which rests on the assumption that the absence of a causal effect has been established.

## Appendix 5

### MeSH disease areas

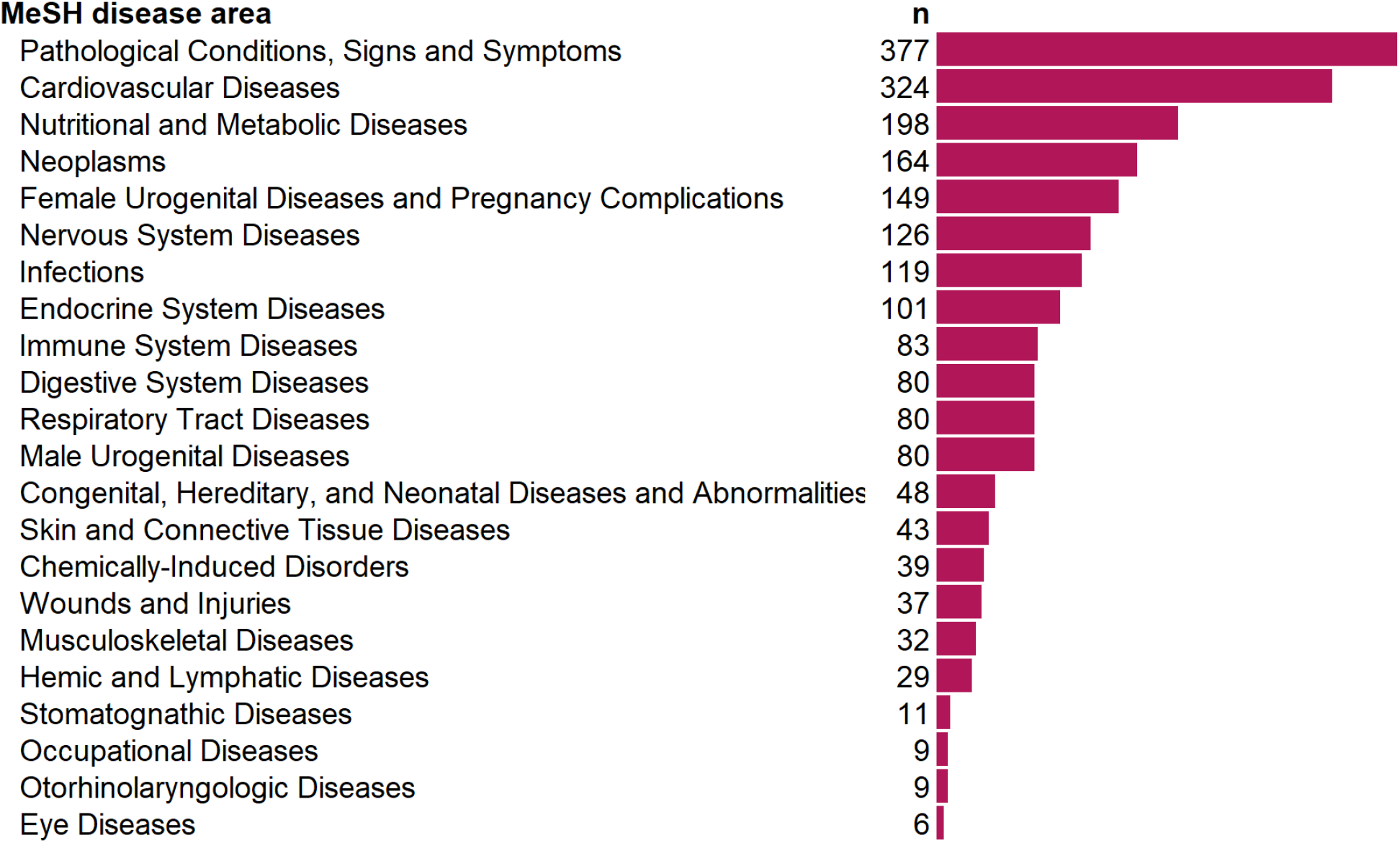

## Appendix 6

### Causal strength over time

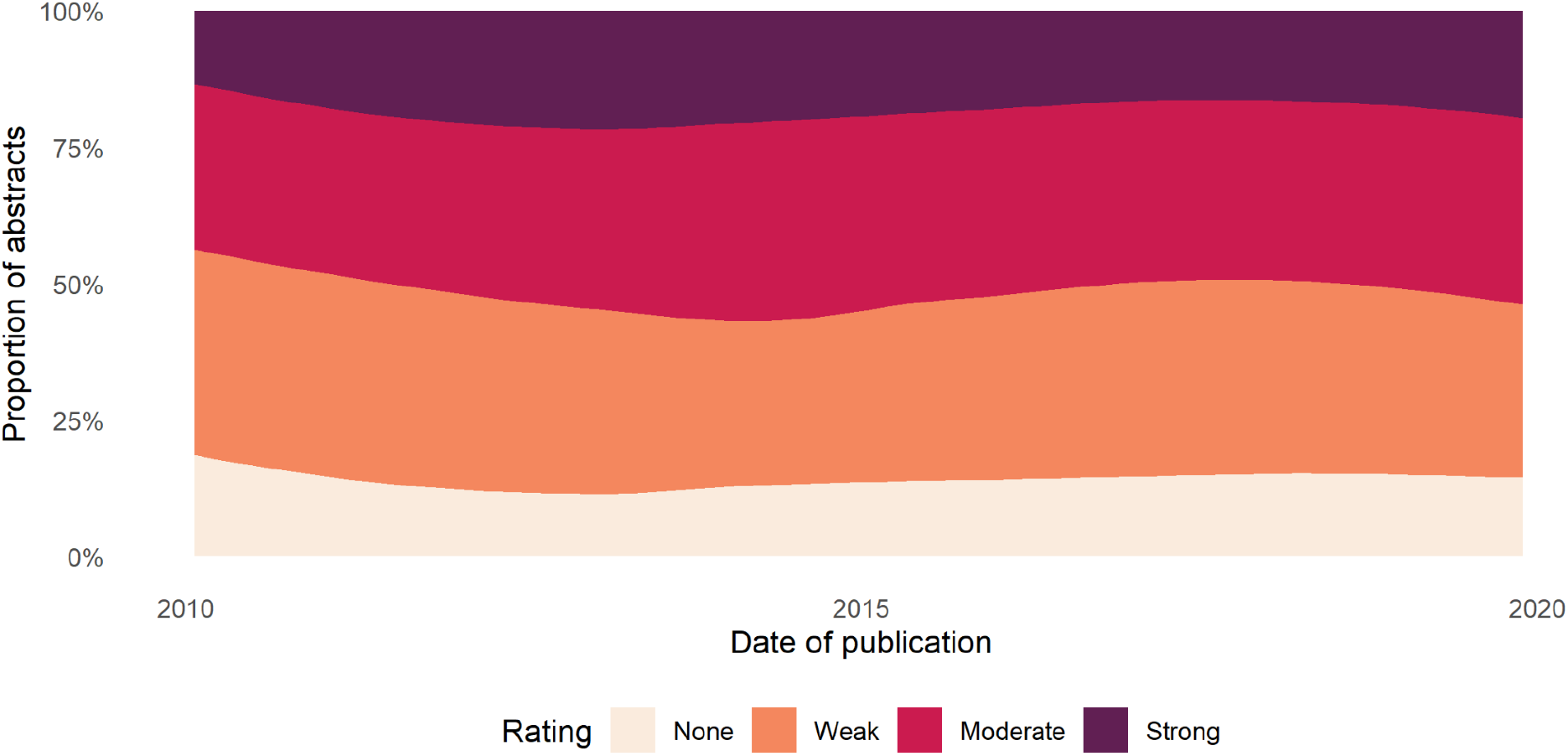

Chart is generated through LOESS smoothing the proportions in each category over time.

## Appendix 7

### Modifying phrases

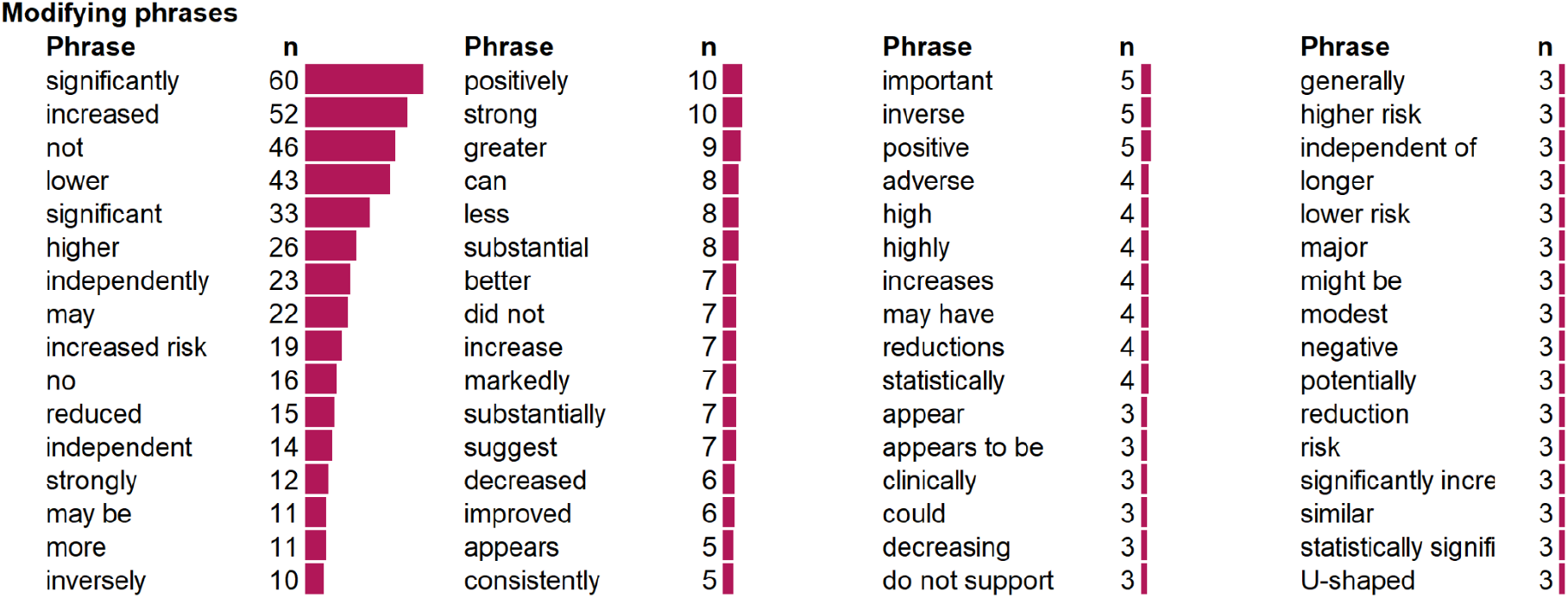

## Appendix 8

### Causal strength of linking sentences in abstract, by journal

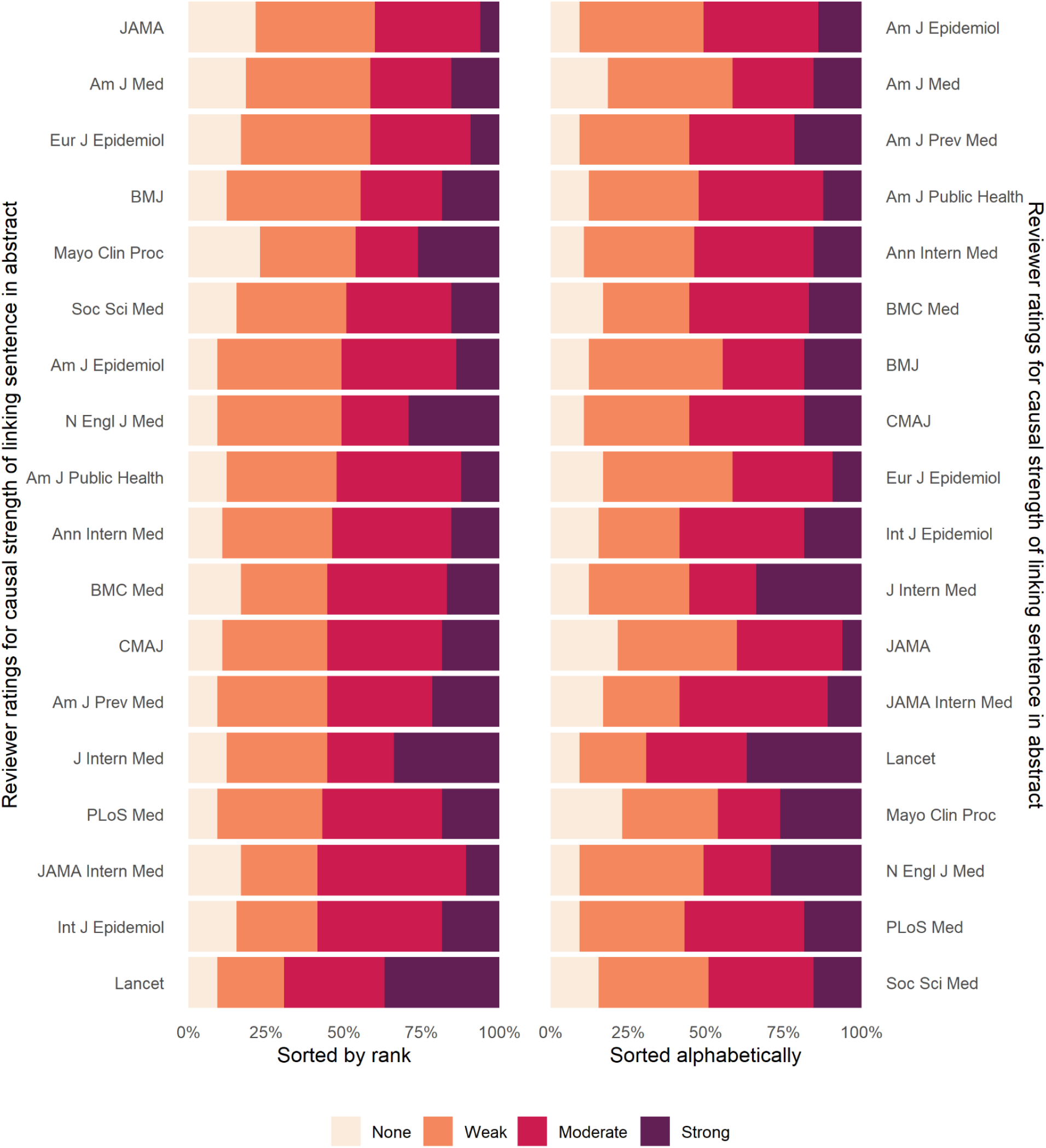

## Appendix 9

### Causal strength action recommendations in abstract, by journal

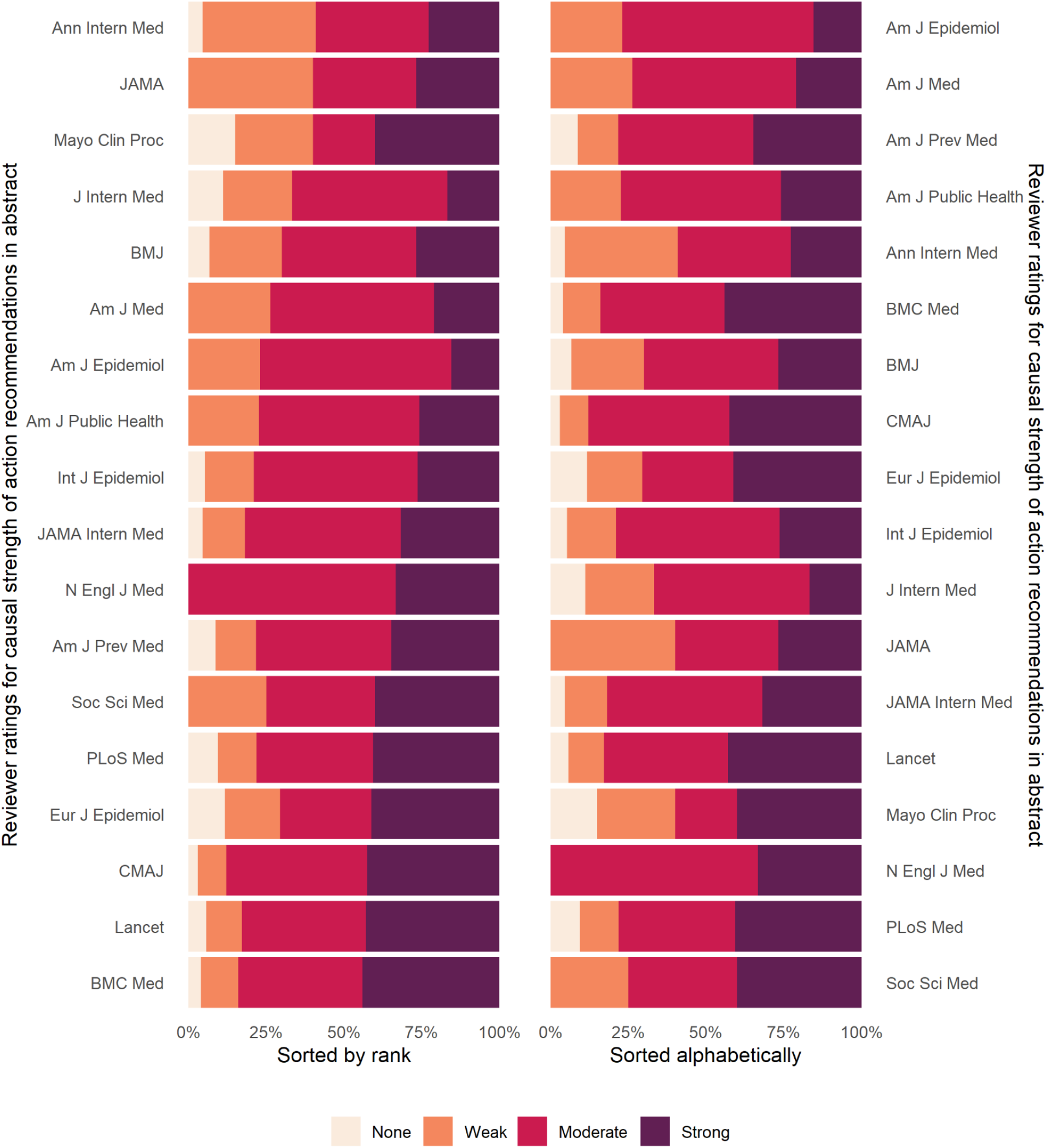

## Appendix 10

### Causal strength action recommendations in abstract including NAs, by journal

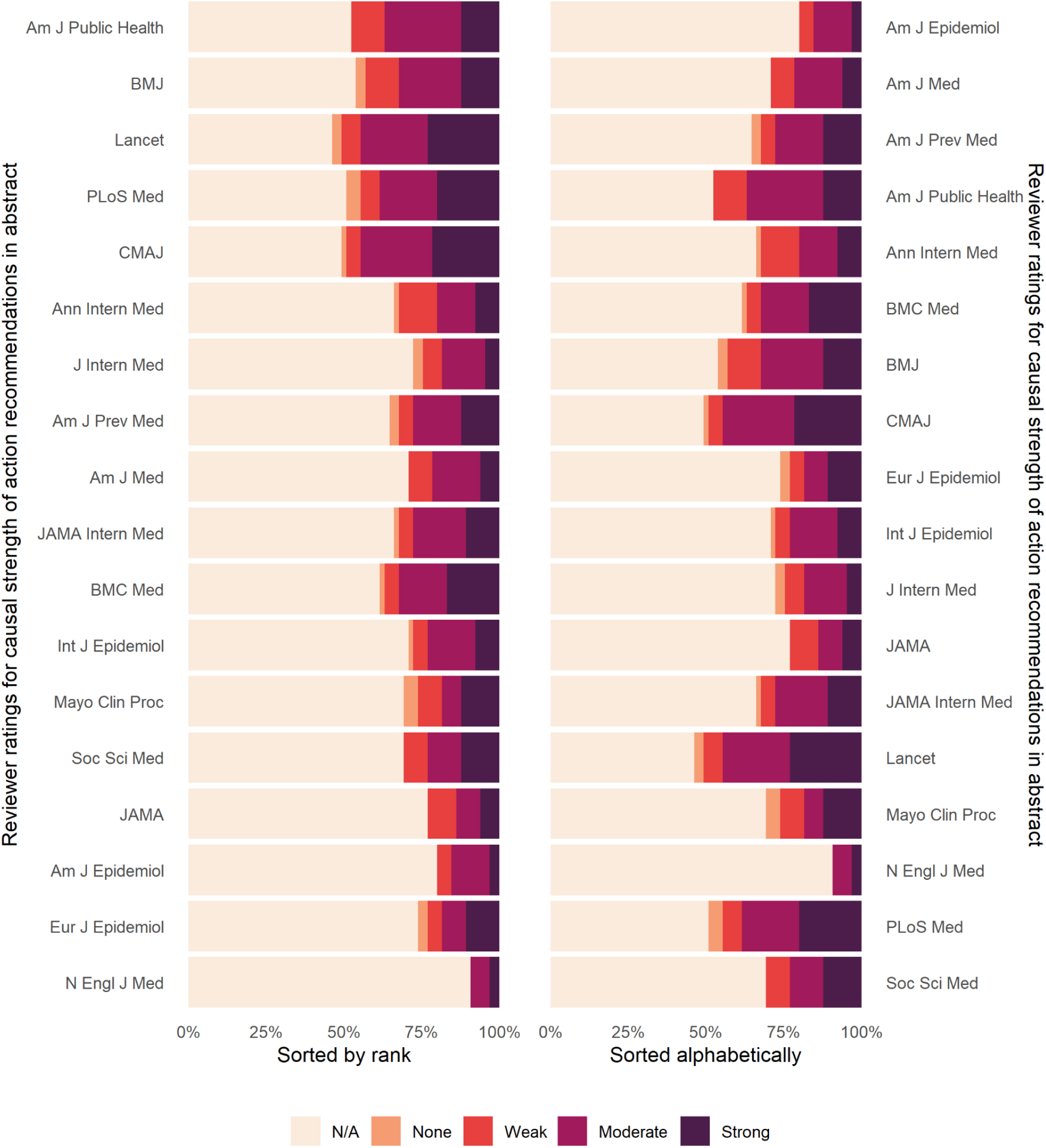

