## Supplementary material for "Causal and Associational Language in Observational Health Research: A systematic evaluation": Review tool (from another version of this preprint: version 4, 2021-10-08)

### Causal language review tool

Version 1.0

* Required

#### Article and reviewer information

Please notify Noah Haber or Sarah Wieten via Slack if something here does not appear correct

1. Record information *

   DO NOT MODIFY

   _______________________________________________
   _______________________________________________
   _______________________________________________
   _______________________________________________
   _______________________________________________

2. Record identifier *

   DO NOT MODIFY

   _______________________________________________

3. Do you wish to recuse yourself from reviewing this study for any reason (e.g. social or professional relationship with the authors, financial conflict of interest, etc)? *

   Recusal will result in reassignment of this review to another reviewer/

   *Mark only one oval.*

   - ( ) No, I do not wish to recuse myself.
   - ( ) Yes, I recuse myself from reviewing this paper. *Skip to question 35*

Inclusion/exclusion criteria

#### Inclusion/exclusion criteria

-Observations must be human- or aggregate group of humans-unit level of observation
The primary research question of the study must be to examine the causal and/or non-causal association between one primary exposure concept and one primary outcome concept
-One "primary" exposure/outcome can include multiple measures of the same or similar broad exposure and/or outcome concept.
-Articles can include many exposures/outcomes, but focus in particular on one exposure/outcome pair as their primary association of interest (e.g. in the title, in the study aims, etc)
-Articles that are about more than one primary concept (e.g. searching for what risk factors are associated with the outcome) will be excluded.
-The primary research question must be examined quantitatively using primary data (i.e,. not a review, meta-analysis)

4. Inclusion/exclusion criteria review request *

   If you believe that this article does not meet the inclusion/exclusion criteria for the Causal Language Study, you may request that the administrators reconsider it for inclusion. If you believe that it does not meet criteria, please explain why in the commeonts box at the end. The administrators will consider inclusion for all requests, and may either decide to remove it from the sample, or instruct you to continue with the review.

   *Mark only one oval.*

   - ( ) I believe that this study meets the inclusion/exclusion criteria
   - ( ) I request that the administrators review this study for inclusion/exclusion criteria *Skip to question 35*

Definitions and guidance for answering questions:

Exposure: For this project, "Exposure" refers to the independent variable of interest (in a regression sense) or the primary or antecedent variable being investigated for a possible (non-)causal link to the study outcome, or resulting or end-point variable. It may be labelled by terms such as treatment, factor, risk factor, protective factor, determinant, intervention, correlate, predictor, agent, cause, causative agent, or other terms.

Outcome: For this project, "Outcome" refers to the dependent or effect variable of interest that is being investigated for a possible link to the exposure (surrogate measures or clinical events). It is typically a post-exposure variable i.e. assumed or known to be preceded by the exposure. It is sometimes called the study endpoint variable, consequence, result, or other terms.

Linking word/phrase: A linking word/phrase describes the nature of the connection between some defined exposure and some defined outcome, generally used in a sentence containing both exposure and outcome. This can describe the type of relationship (e.g. "associated with") and/or differences in levels (e.g. "had higher") that may or may not be causal in nature. For our purposes, the phrase may contain 1-3 words, where one of the words is a preposition to link the exposure and outcome. Some examples may include constructions such as "associated with," "effect of," "increased," "was higher than," "correlated with," "caused," "harms," "predicts," "risk factor for," "determined," "impacts," "decreased," "linked to," etc.

Modifying word/phrase: A modifying word/phrase is a word or phrase that modifies the linking word/phrase describing the nature of the relationship between the exposure and outcome. This includes adding signals of direction, strength, doubt, negation, and statistical properties to the relationship. This may include phrases like "may be," "positively," "strongly", "potentially", "is likely to," "does/is not," "statistically significant," etc.

Causal language: Causal language implies that one entity influences (or does not influence) another. We define language as being causal if that language implies that movement (or lack thereof) in the outcome was either 1) impelled by the exposure of interest (i.e. a change in the exposure drives or does not drive a change in the outcome, e.g., increase, decrease, improve, change), or 2) implies attribution of the outcome to the exposure (i.e. assigns the responsibility for the change or lack of change in the outcome to the exposure, e.g. "due to," "since," "attributable to").

Action recommendation: This is a description of how a consumer of the research in question might utilize the results and conclusions of the research. This may include recommending that some actor consider changes (or no changes) in some set of procedures and actions. Action recommendations concern what to do with the research. For our purposes, we do not count calls for additional research as action recommendations.

Causal implication of recommendations: Recommendations may often imply a causal interpretation of a finding. For example, authors may suggest that it could be beneficial to change the amount of an exposure, which rests on the assumption that the exposure has a causal effect on the outcome. As a variation, it may also be suggested that an exposure need not be changed, which rests on the assumption that the absence of a causal effect has been established.

5. Title/Abstract: Primary exposure of interest *

A few word description, copied directly from the title and/or abstract. Gather from the title if available, and if not the introduction section, discussion section, or results section of the abstract (in that order).

____________________

6. Title/Abstract: Primary outcome of interest *

A few word description, copied directly from the title and/or abstract. Gather from the title if available, and if not the introduction section, discussion section, or results section of the abstract (in that order).

____________________

#### Abstract: Linking sentence section

7. Abstract: Primary linking sentence(s) *

What is the primary sentence / phrase used that contains the linking phrase between the exposure and outcome? Copy and paste from the abstract. Preference for this sentence if there are multiple potential sentences should be in order of 1) the conclusions section of the abstract (if available), 2) the discussion section of the abstract, 3) the results section of the abstract (if available), or 4) somewhere else (or unlabelled sections). Search in particular for a sentence which contains the exposure, outcome, linking word, and any modifying phrases. If more than one sentence is necessary, copy all necessary sentences. If there are multiple sentences that equally meet these guidelines, choose the first one to occur.

____________________

____________________

____________________

____________________

____________________

8. Abstract: Primary linking sentence location *

Where is the primary sentence / phrase used that contains the linking phrase between the exposure and outcome located?

*Mark only one oval.*

- ◯ Abstract conclusions section
- ◯ Abstract discussion section
- ◯ Abstract results section
- ◯ This abstract contains no structured sections.
- ◯ Other: ____________________

9. Abstract: Primary linking word/phrase *

Based on the sentence copied above, select the primary linking word/phrase from the above sentence and copy it here. The word or phrase should be up to three words maximum, often including a preposition. It should not contain any modifying phrases. If there are multiple words/phrases that equally meet these guidelines, choose the first one.

____________________

10. Abstract: Modifying word/phrases

Based on the sentence above, copy any modifying words or phrases that modify the nature (e.g. strength, intensity, room for doubt, negation, direction, etc.) of the primary linking word/phrase from the above sentence and paste them here. If not, leave this blank. If multiple, separate with a semicolon.

____________________

11. Abstract: How strongly does the language in this sentence imply that the authors identified a causal relationship between the primary exposure and the primary outcome? *

Based on the linking sentence copied above, how strongly do you believe that this linking sentence implies a causal relationship between the exposure and outcome?

*Mark only one oval.*

- None: The linking sentence does not imply in any way a causal relationship was identified.
- Weak: The linking sentence might imply a causal relationship was identified, but it is unclear or possible to come to that conclusion in the absence of any causal inference.
- Moderate: The linking sentence mostly implies a causal relationship was identified, but it is unclear or possible to come to that conclusion in the absence of any causal inference.
- Strong: The linking sentence clearly implies that causality had been identified.

**Abstract: Action recommendation(s) section**

12. Abstract: Action recommendation(s)

Copy (if any) what major claims are made about how a consumer of this research might utilize its results and conclusions. If there are multiple, choose the one which maximally implies causal inference in the following question. Note: Actions calling for more research do not apply here. If none, leave blank.

13. Abstract: Action recommendation causal implication *

Choosing the claim which most implies or requires that the evidence from this study was causal in nature, how strongly is this implication made?

*Mark only one oval.*

- N/A: No action recommendation(s) provided in this abstract.
- None: The action recommendation would be made appropriately in the absence of any causal relationship.
- Weak: The action recommendation may be made appropriately had a causal relationship been identified, but it is unclear or possible to come to that recommendation in the absence of any causal inference.
- Moderate: The action recommendation most likely could only be made appropriately had a causal relationship been identified, but it is unclear or possible to come to that recommendation in the absence of any causal inference.
- Strong: The action recommendation could only be made appropriately had a causal relationship been identified.

14. Skip pattern marker

DO NOT MODIFY

*Mark only one oval.*

- TA *Skip to question 35*
- FT

Full text: Discussion/conclusions

The following questions should come from the discussion and/or conclusions section or the equivalent.

**Discussion/conclusions: Linking sentence section**

15. Discussion/conclusions: Primary linking sentence(s) *

What is the primary sentence / phrase used that contains the linking phrase between the exposure and outcome? Copy and paste from the discussion or conclusions section. Preference for this sentence if there are multiple potential sentences should be in order 1) the first paragraph of the discussion or conclusions section, 2) elsewhere in the discussion or conclusions section, 3) results section, 4) elsewhere in the text. Search in particular for a sentence which contains the exposure, outcome, linking word, and any modifying phrases. If more than one sentence is necessary, copy all necessary sentences. If there are multiple sentences that equally meet these guidelines, choose the first one.

16. Discussion/conclusions: Primary linking sentence location *

Where is the primary sentence / phrase used that contains the linking phrase between the exposure and outcome located?

*Mark only one oval.*

- First paragraph of the discussion or conclusions section
- Elsewhere in the discussion or conclusions section
- Results section
- Other: ______________________

17. Discussion/conclusions: Primary linking word/phrase *

Based on the sentence copied above, select the primary linking word/phrase from the above sentence and copy it here. The word or phrase should be up to three words maximum, often including a preposition. It should not contain any modifying phrases. If there are multiple words/phrases that equally meet these guidelines, choose the first one.

______________________

18. Discussion/conclusions: Modifying word/phrases

Based on the sentence above, copy any modifying words or phrases that modify the nature (e.g. strength, intensity, room for doubt, negation, direction, etc.) of the primary linking word/phrase from the above sentence and paste them here. If not, leave this blank. If multiple, separate with a semicolon.

______________________

19. Discussion/conclusions: How strongly does the language in this sentence imply that the authors identified a causal relationship between the primary exposure and the primary outcome? *

Based on the linking sentence copied above, how strongly do you believe that this linking sentence implies a causal relationship between the exposure and outcome?

*Mark only one oval.*

- None: The linking sentence does not imply in any way a causal relationship was identified.
- Weak: The linking sentence might imply a causal relationship was identified, but it is unclear or possible to come to that conclusion in the absence of any causal inference.
- Moderate: The linking sentence mostly implies a causal relationship was identified, but it is unclear or possible to come to that conclusion in the absence of any causal inference.
- Strong: The linking sentence clearly implies that causality had been identified.

**Discussion/conclusions: Action recommendation(s) section**

20. Discussion/conclusions: Action recommendations(s)

Copy (if any) what major action recommendation claims made about how to utilize the evidence from this research. If there are multiple, choose the one which maximally implies causal inference in the following question. Note: Actions calling for more research do not apply here. If none, leave blank.

______________________

21. Discussion/conclusions: Action recommendation causal implication *

Choosing the claim which most implies or requires that the evidence from this study was causal in nature, how strongly is this implication made?

*Mark only one oval.*

- N/A: No action recommendation(s) provided.
- None: The action recommendation would be made appropriately in the absence of any causal relationship.
- Weak: The action recommendation may be made appropriately had a causal relationship been identified, but it is unclear or possible to come to that recommendation in the absence of any causal inference.
- Moderate: The action recommendation most likely could only be made appropriately had a causal relationship been identified, but it is unclear or possible to come to that recommendation in the absence of any causal inference.
- Strong: The action recommendation could only be made appropriately had a causal relationship been identified.

**Discussion/conclusions: Miscellaneous**

22. Discussion/conclusions: Causal disclaimer statements

If there are any statements in the discussion which explicitly mention causality as a cautionary or disclaimer statement, paste them here. Examples may include "correlation does not imply causation" or similar. If not, leave blank.

#### Full text: Introduction/Methods

The following questions should come from the introduction and/or methods section or the equivalent.

23. Introduction: Causal theory explanation sentence in introduction

Copy sentence(s) in the introduction section which contain informal theoretical discussion for why the exposure might be causally linked to the outcome of interest. If no such sentence exists, leave blank.

24. Introduction: Theory explanation causal implication strength *

How strongly does the above sentence(s) imply that there is a causal relationship of interest?

*Mark only one oval.*

- N/A: No discussion of how the exposure and outcome are linked is provided.
- None: No theory discussion is provided which implies a possible causal relationship between the outcome and exposure.
- Weak: The introduction contains theory which may imply a causal relationship, but it is unclear or ambigous..
- Moderate: The introduction contains theory which implies a causal relationship, but with some plausible room for doubt.
- Strong: The introduction contains theory about the relationship between exposure and outcome that is explicitly and/or unambiguously causal.

25. Introduction / Methods: Formal causal model *

Is any formal causal model presented anywhere in this paper? That might include a graphical causal model, equations, simulations, etc. While this question is in the Introduction/Methods section of this review tool, please include any formal causal model found anywhere in the article.

*Check all that apply.*

- [ ] Causal Directed Acyclic Graph (DAG)
- [ ] Other graphical causal model (describe in "other")
- [ ] Structural equations model
- [ ] Equation-based toy model
- [ ] Simulation model
- [ ] No, there are no formal causal models presented in this paper.

Other: [ ] ____________________

26. Introduction / Methods: Are there variables controlled, adjusted, matched, or stratified on? *

This should be for the "main" specification only (i.e. the result most prominently displayed in the abstract)

*Mark only one oval.*

- No (0)
- Yes (1-3 variables)
- Yes (4-9 variables)
- Yes (10+ variables)

27. Anywhere in text: Is "confounding" or "confounders" discussed or mentioned in relation to the methods, results, and/or interpretation of this study? Please check all that apply. *

The specific word "confound*" must appear, where * can be any suffix. This must be in reference to the methods, results, and/or interpretation of this study, and not in reference to other studies.

*Check all that apply.*

- [ ] No, this does not appear in the study manuscript
- [ ] Yes, in the introduction
- [ ] Yes, in the methods section
- [ ] Yes, in the discussion limitations section
- [ ] Yes, elsewhere in the discussion section

Other: [ ] ______________________

28. Does the article contain a pop-out section? *

This section only pertains to studies which have "pop out" sections separate from the main text that summarize, explain, and/or translate the information in the study. If there are no pop out sections, skip the rest of these questions.

*Mark only one oval.*

( ) Yes
( ) No *Skip to question 35*

Full text: Pop-out

This section only pertains to studies which have "pop out" sections separate from the main text that summarize, explain, and/or translate the information in the study. If there are no pop out sections, skip the rest of these questions

**Pop-out: Linking sentence section**

29. Pop-out: Primary linking sentence(s)

What is the primary sentence / phrase used that contains the linking phrase between the exposure and outcome? Copy and paste here. Search in particular for a sentence which contains the exposure, outcome, linking word, and any modifying phrases. If more than one sentence is necessary, copy all necessary sentences. If there are multiple sentences that equally meet these guidelines, choose the first one.

______________________

30. Pop-out: Primary linking word/phrase

Based on the sentence copied above, select the primary linking word/phrase from the above sentence and copy it here. The word or phrase should be up to three words maximum, often including a preposition. It should not contain any modifying phrases. If there are multiple words/phrases that equally meet these guidelines, choose the first one.

______________________

31. Pop-out: Modifying word/phrases

Based on the sentence above, copy any modifying words or phrases that modify the nature (e.g. strength, intensity, room for doubt, negation, direction, etc.) of the primary linking word/phrase from the above sentence and paste them here. If not, leave this blank. If multiple, separate with a semicolon.

______________________

32. Pop-out: How strongly does the language in this sentence imply that the authors identified a causal relationship between the primary exposure and the primary outcome? *

Based on the linking sentence copied above, how strongly do you believe that this linking sentence implies a causal relationship between the exposure and outcome?

*Mark only one oval.*

( ) None: The linking sentence does not imply in any way a causal relationship was identified.
( ) Weak: The sentence might imply a causal relationship was identified, but it is unclear or possible to come to that conclusion in the absence of any causal inference.
( ) Moderate: The sentence mostly implies a causal relationship was identified, but it is unclear or possible to come to that conclusion in the absence of any causal inference.
( ) Strong: The sentence clearly implies that causality had been identified.

**Pop-out: Action recommendation(s) section**

33. Pop-out: Action recommendations(s)

Copy (if any) what major action recommendation claims made about how to utilize the evidence from this research. If there are multiple, choose the one which maximally implies causal inference in the following question. Note: Actions calling for more research do not apply here. If none, leave blank.

34. Pop-out: Action recommendation causal implication

Choosing the claim which most implies or requires that the evidence from this study was causal in nature, how strongly is this implication?

*Mark only one oval.*

- N/A: No action recommendation(s) provided
- None: The action recommendation would be made appropriately in the absence of any causal relationship.
- Weak: The action recommendation may be made appropriately had a causal relationship been identified, but it is unclear or possible to come to that recommendation in the absence of any causal inference.
- Moderate: The action recommendation most likely could only be made appropriately had a causal relationship been identified, but it is unclear or possible to come to that recommendation in the absence of any causal inference.
- Strong: The action recommendation could only be made appropriately had a causal relationship been identified.

Submit form

35. Additional comments

This field is for comments intended to communicate anything additional you would like to the arbitrators and/or the public about this study. These comments will be made public as part of the study dataset. If you have general questions or comments and/or would like your comments to be private, please use the Slack instead.

36. Check here to confirm that you have filled out the above information to the best of your abilities *

To complete submission, you must check this button.

*Check all that apply.*

- [ ] I confirm

This content is neither created nor endorsed by Google.

Google Forms
